# Cardiovascular and cancer mortality inversely associated with long-term ambient ultraviolet exposure across US Health Service Areas with UK Biobank corroboration

**DOI:** 10.64898/2026.10.06.26364908

**Authors:** J MacMahon, C Emig, PH Hart, B Wang, R Weller, C Dibben, A Stevenson

## Abstract

*OBJECTIVE:* To assess associations of ambient UV with cardiovascular and cancer mortality across US Health Service Areas (HSAs), prioritise outcomes by signal and attributable burden, assess racial and ethnic variation, with UK Biobank corroboration.

*DESIGN:* Ecological analysis of HSAs using linear regression of crude mortality on ambient UV and social vulnerability, with correction for multiple comparisons.

*SETTING:* US HSAs and UK Biobank.

*DATA SOURCES:* Publicly available US data and UK Biobank data accessed under Application 82526.

*PARTICIPANTS:* 704 808 annual cardiovascular deaths (aged ≥65) and 565 287 annual cancer deaths (aged ≥50) from populations of 52.8 and 115.2 million; UK Biobank analyses included 412 665 individuals.

*MAIN OUTCOME MEASURES:* Cardiovascular, all-cancer, and body-site-specific cancer mortality; regression slope per interquartile-range increase in UV (β UV/IQR), modelled attributable burden for US data, and hazard ratios for the UK Biobank data.

*RESULTS:* Higher UV was associated with lower cardiovascular (βUV/IQR −68.3 deaths/100 000; P<0.001) and all-cancer mortality (−43.2; P<0.001), corresponding to 36 043 (5.1%) and 49 789 (8.8%) fewer US deaths annually, respectively. Associations varied by race and ethnicity. Cardiovascular findings were directionally consistent between US and UK datasets. All-cancer mortality and 10 of 18 cancers were inversely associated with UV in the US; UK analyses found inverse associations for all-cancer and two individual body-site-specific cancers. Twelve cancers showed directionally concordant associations across the datasets, of which ten were inverse. Lung cancer showed the strongest inverse association in each dataset.

*CONCLUSIONS:* Higher UV was associated with lower cardiovascular and cancer mortality across US HSAs, with modelled attributable burdens exceeding 5%; UK Biobank findings provided partial corroboration. Twelve of 18 site-specific cancers were directionally concordant between datasets. US race-stratified heterogeneity did not follow a skin-colour gradient, suggesting residual confounding.

*WHAT IS ALREADY KNOWN ON THIS TOPIC:* Although intermittent ultraviolet (UV) exposure is associated with risk of increased skin-cancer incidence including keratinocyte cancers and melanoma, there is no parallel increase in mortality. In contrast, long-term ecological and cohort studies have consistently associated higher UV exposure and sun-seeking behaviour with lower all-cause, cardiovascular, and internal cancer mortality. While large-scale randomised placebo-controlled trials of vitamin D supplements have all failed their primary endpoints to reduce incidence or mortality, shifting attention toward other UV-induced pathways that may better explain the observed net benefits. Complementary clinical evidence comes from NB-UVB phototherapy, a controlled surrogate of solar UV, which provides mechanistic insight through its association with reduced cardiovascular and cerebrovascular events and its guideline-directed role in dermatological conditions and early-stage cutaneous T-cell lymphoma.

*What this study adds:* This study provides updated nationwide estimates of the association between long-term ambient ultraviolet radiation and mortality across US Health Service Area and UK Biobank datasets, showing inverse associations with two of the leading causes of death: cardiovascular disease and cancer. Across the observed US interquartile range of ambient UV, higher UV was associated with 36,043 fewer cardiovascular deaths and 49,789 fewer cancer deaths per year, with inverse cancer associations observed across multiple internal cancers and directionally corroborated in an independent UK Biobank dataset across all-cancer and 12 of 18 individual site-cancers. Prespecified racial subgroup analyses showed that UV–mortality relationships were heterogeneous and non-monotonic across racial groups, rather than following the expected gradient by skin colour seen in clinical phototherapy. This suggests that UV–mortality associations may be shaped by additional biological, behavioural, environmental, or social determinants not fully captured by the Social Vulnerability Index.

## Introduction

For more than a century, observational work has suggested that ambient sunlight and UV (ultraviolet) radiation exposure are inversely correlated with several internal cancer outcomes, including incidence and survival, even while increasing the incidence of skin cancer.^1^ More recent analyses have repeatedly documented inverse correlations between UV-related measures and cancer mortality, raising the possibility of systemic benefits beyond the skin.^2^ Vitamin D, produced by the skin under UV exposure, was the presumed mechanism, with serum vitamin D the biomarker surrogate.^3^

Following these strong observational associations between vitamin D status and both cardiovascular and cancer mortality, multiple large-scale trials were initiated. Although low 25(OH)D levels are associated with cardiovascular and cancer mortality, none of the randomised placebo-controlled trials demonstrated a statistically significant benefit of vitamin D supplementation for a prespecified primary cardiovascular, cancer, mortality, or survival endpoint.^4–8^ An individual-participant meta-analysis of 104,727 participants likewise found no significant overall effect on cancer mortality (RR 0.94, 95% CI 0.86 to 1.02). Neither preventive nor therapeutic trials of vitamin D supplementation have reproduced the observational UV-mortality associations (Supplementary Table S1), which supports investigation of vitamin D-independent pathways.^9,10^

Contrary to the assumption that Vitamin D supplementation impacts the immune system the same as the UV spectrum of sunlight head-to-head trial of vitamin D supplements and NB-UVB resulted in a differential immune response. In a randomised trial, Ponda et al. repleted vitamin D–deficient adults using either oral vitamin D₃ or NB-UVB. Despite comparable improvement in 25(OH)D, peripheral-blood gene-set enrichment diverged: interferon-α and interferon-γ responses were positively enriched after oral vitamin D₃ but negatively enriched after NB-UVB.^11^

The prospective Melanoma in Southern Sweden (MISS) cohort of over 30,000 women followed for a mean of 25 years was established to confirm the harms of sun-seeking. However, it provides the strongest evidence that the net benefit of sun-seeking exceeds the melanoma risk, with the most active sun-seeking habits correlating with the lowest all-cause mortality and the longest life expectancy. The analyses showed that much of the survival advantage arose from decreased cardiovascular and noncancer deaths. A later analysis of the same cohort separated consistent solarium use from intermittent behaviour: both groups had approximately 18% lower all-cause mortality than non-users, with no difference in melanoma-specific mortality, while melanoma risk was increased only among solarium users with reported intermittent solar overexposure. (HR 1.60, 95% CI 1.30 to 2.00).The pattern that recurs across these subsets of the MISS cohort is therefore a divergence between incidence and mortality: intermittent overexposure raises the incidence of melanoma and keratinocyte cancers without a parallel increase in mortality.^12–14^

These findings have since been reproduced in other large studies, including our analyses of the UK Biobank, where higher ambient UV radiation was associated with lower all-cause and cardiovascular mortality.^15,16^ Our analyses included 395,086 participants of white European ancestry and approximated both active UV exposure (solarium use) and passive exposure (annual average ambient residential shortwave radiation at the primary residence). Both groups showed improved survival from all-cause, cardiovascular and cancer mortality.

Prospective, observational, and population-based UV research signals of inversed associations with cardiovascular and cancer mortality are consistent but may be confounded by lifestyle.^2,9,12–15^ No US study has yet quantified the association between ambient UV and cause-specific mortality at national population scale while accounting for area-level social vulnerability and across racial groups. We therefore used an ecological Health Service Area design that captured population-level ambient UV levels to quantify associations between long-term ambient UV and cardiovascular and cancer mortality. We prioritised outcomes by signal magnitude and attributable burden, assessed variation by race/ethnicity, and corroborated the findings against a UK Biobank cohort.

## Methods

### All the US environmental and mortality input data are from publicly available sources. The UK Biobank cancer and mortality data are described below and in appendix E Ambient Ultraviolet Radiation

The county-level ambient UV data were sourced from the National Cancer Institute (NCI) Geographic Information Systems web portal,^17^ and provided historical Average Daily Global Solar Radiation estimates (AVGLO), a proxy measure for UV, in Wh/m² in 5-year annual averages for years 2000-2024.18 The average of these 5-year averages across 2000-2024 was chosen as the longest annual data set available for the smoothest accumulated signal in which a consistent algorithm overlapped the mortality data set. AVGLO represents broadband global solar radiation and is provided through the NCI GIS portal as a validated geographic proxy for ambient UV exposure in population-based cancer research.

**Figure 1.**
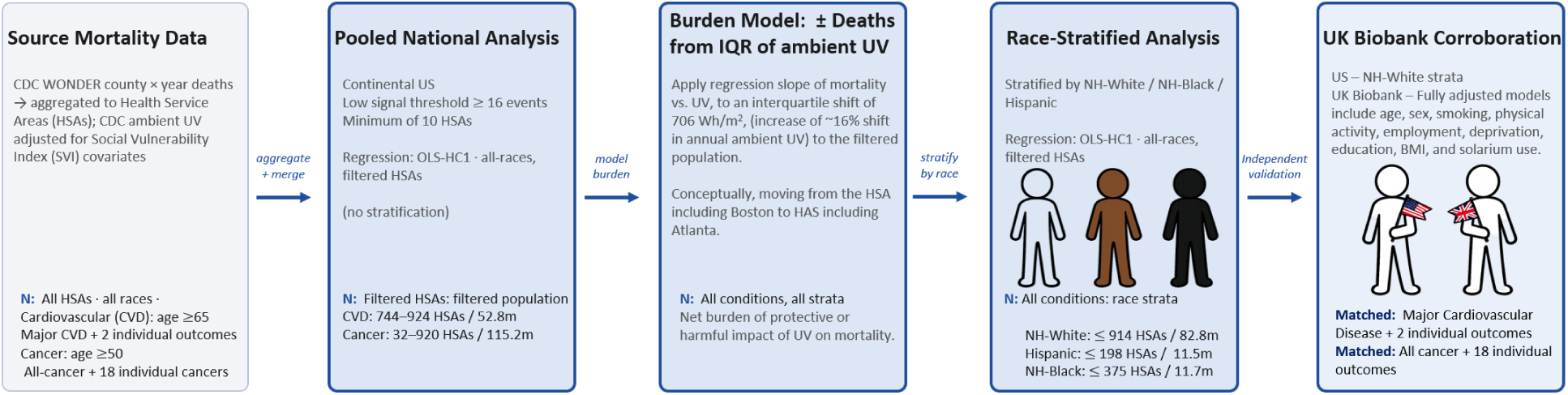
Process for generating the national-scale UV versus mortality analysis, deriving an association signal, converting that signal into a modelled mortality burden, and corroboration with the UK Biobank.^15^

### Social Vulnerability Adjustment

The 2018 CDC/ATSDR Social Vulnerability Index (SVI) served as the area-level covariate for socioeconomic and demographic deprivation, taken at county level and combining 15 American Community Survey indicators across four themes.^19^ Its empirical validation as a predictor of population mortality across both endpoint families is summarised in Supplementary Appendix C.

**Figure 2.**
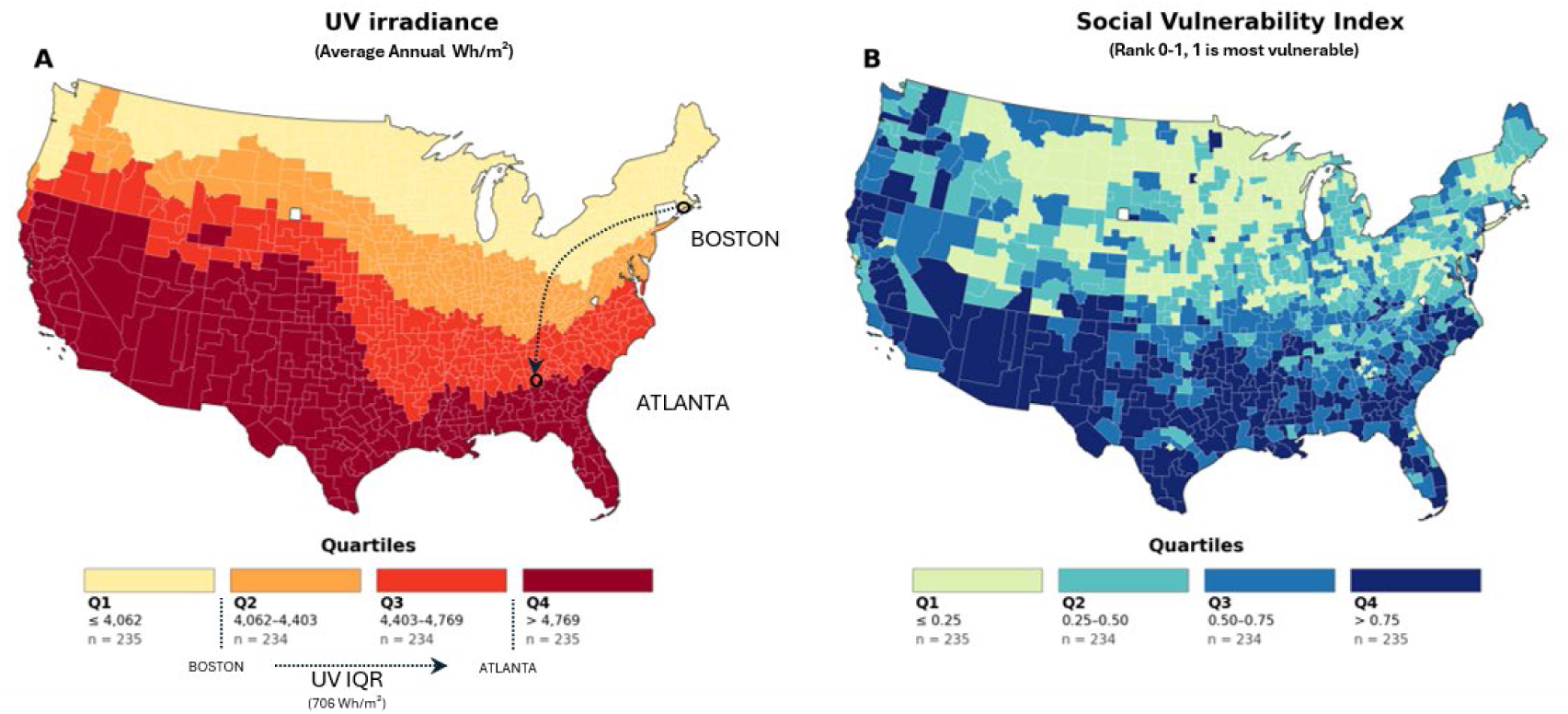
Geographic distribution of ultraviolet irradiance and social vulnerability across 937 Health Service Areas in the contiguous United States, before outcome-specific stability and complete-case filters were applied. (A) Mean annual ultraviolet irradiance during 2000-2024, categorised into quartiles. Boston and Atlanta illustrate an approximate interquartile-range contrast of 706 Wh/m².^15,17^ (B) CDC 2018 Social Vulnerability Index overall percentile rank, categorised into quartiles; higher ranks indicate greater social vulnerability.^19^, Values are population-weighted to the HSA level. HSA, Health Service Area; IQR, interquartile range; SVI, Social Vulnerability Index; UV, ultraviolet.

### Mortality Data

County-level underlying-cause mortality counts and populations for 2018-2019 were obtained from the Centers for Disease Control and Prevention’s Wide-ranging Online Data for Epidemiologic Research (WONDER) database.^20^ The two pre-COVID years were combined to improve estimate stability. Data were obtained in 5-year age groups and restricted to adults aged ≥65 years for cardiovascular outcomes and ≥50 years for cancer outcomes. Cancer sites were selected from the NCI State Cancer Profiles priority endpoints.^21^ Endpoint definitions and ICD-10 codes are from the CDC WONDER request interface and provided in Supplementary Table S2.^22^

### Aggregation to Health Service Areas

Counties were assigned to NCI-modified Health Service Areas (HSAs) using the SEER County Attributes crosswalk.^23^ HSAs group contiguous counties into hospital-care markets, providing an intermediate geographic scale between 3,000+ counties and 48 contiguous states that preserves regional UV variation while improving the stability of mortality estimates. HSAs have been used in US small-area mortality research and are directly compatible with the county-level WONDER, NCI UV, and CDC/ATSDR SVI inputs used here.^24^

County UV and SVI values were population-weighted to the HSA level, deaths and populations were summed across constituent counties and years, and crude rates were calculated from those totals. Suppression handling and the stability filters are detailed in Supplementary Appendix B, and the derivation of the analytic sample is shown in Supplementary Figure S1. The primary cardiovascular and cancer datasets included 930 and 920 HSAs, representing 52.8 million adults aged 65 years or older and 115.2 million adults aged 50 years or older, respectively.

### The Signal Analysis

OLS regressions of HSA-level average annual crude mortality rates (deaths per 100,000 person-years) were fitted on UV and SVI, mutually adjusted, with HC1 standard errors^25^ for the heterogeneity in HSA population sizes. UV and SVI co-vary across US geography, so both terms were required in the model to isolate the UV signal. Linearity of the UV-mortality relationship was examined in supplementary nonlinear exposure-response analyses. Univariate UV and SVI models were fitted in parallel and are reported in Supplementary Table S3 and Supplementary Figure S3 (reciprocal UV and SVI adjustment).

Mortality was modelled as a crude rate within age-restricted populations (cancer 50+, cardiovascular 65+), not age-standardised; an age-adjustment sensitivity analysis is reported in the appendix. The age floors follow SEER/NCI and CDC/AHA reporting conventions and concentrate the analysis on the age segments carrying the bulk of the mortality.

Benjamini–Hochberg false discovery rate (FDR) correction was applied independently by tier, with the aggregate endpoints tested at q < 0.05 and each individual-outcome tier corrected for its own number of tests.^26^ The tier definitions and resulting thresholds are given in Supplementary Appendix C.

Regression slopes were rescaled from deaths per 100,000 per 1 Wh/m² to deaths per 100,000 per interquartile-range increase in UV, the interquartile range being 706 Wh/m². The derivation is in Supplementary Appendix C.

### Burden Calculation

Modelled mortality burden was calculated as the outcome’s β per UV interquartile range multiplied by its analysed population and divided by 100,000. The full derivation and its terms are given in Supplementary Appendix C. For scale, one interquartile range of 706 Wh/m² is approximately the difference in annual UV between the Health Service Areas centred on Boston, Massachusetts and Atlanta, Georgia.

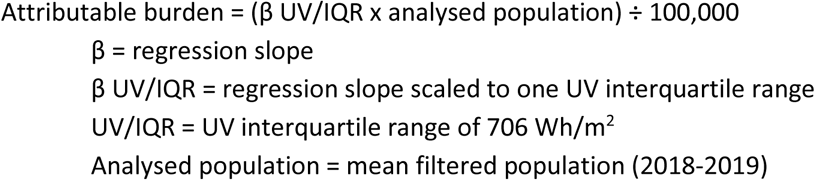

### Race-stratified Assessment

WONDER mortality and populations were downloaded for all races and for the exclusive NH-White, Hispanic and NH-Black strata. The stratum definitions follow WONDER’s Single-Race categories.

Analyses were performed in Python (pandas, numpy, statsmodels OLS with cov_type=’HC1’). The v29 WONDER Mortality Engine pipeline is available from the corresponding author on reasonable request.

### Corroboration Datasets for NH-White stratified Assessment

One historic dataset^2^ and our new expanded UK Biobank data sets provide corroboration to the US results. Both of these are distinct NH-White cohorts, to avoid any pigment/race-based dissonance in UVB response. Boscoe and Schymura compared cancer mortality between northern and southern US states, contrasting erythemally weighted ultraviolet exposure typical of northern Maine, Minnesota or Washington with that typical of southern Florida, Texas or Arizona, an annual average of 650 vs 1,540 kJ/m².^2^

Our UK Biobank analyses identified 412,665 participants of European ancestry and report cardiovascular and cancer mortality. Both datasets are described in Supplementary Appendix E.

### UK Biobank corroboration

Both data sets defined endpoints under the prespecified ICD-10 codes. Both data sets applied Benjamini–Hochberg false discovery rate (FDR) correction for individual cancer analyses.

Complementary analyses in the same 412,665-participant UK Biobank sample includes patient level data and allows for a higher resolution and Cox proportional hazard ratio assessments, including age-adjusted and fully adjusted models. Covariates, the analytic sample definition and its relationship to the previously published complete-case sample are given in Supplementary Appendix E.

### Patient and public involvement

Patients and the public were not involved because this study was a secondary analysis of de-identified, previously collected data.

## Results

### Primary US Analysis

The WONDER database downloads and subsequent filters produced a large cohort for analysis for both major CVD and cancer outcomes of greater than 52.8M people, age ≥ 65, and 115.2M people age ≥ 50, and discrete annual deaths of 704,808 and 565,287 deaths, respectively. Race-stratified baselines were heterogeneous: the NH-White and NH-Black cohorts had above average mortality while the Hispanic cohort was 27% and 38% below the national average for cardiovascular and cancer mortality respectively (Table 1). At the national level, this variance does not follow the skin-pigmentation gradient anticipated from clinical phototherapy which would suggest a melanin based gradient from NH-White, Hispanic, to NH-Black.

**Table 1.** Analytic coverage, annual deaths, crude mortality rates, and ambient ultraviolet exposure by outcome family and race/ethnicity.

| Stratum | HSAs, n | Population (M) | Mean annual deaths | Crude rate /100k | % of All-races | Mean UV (Wh/m<sup>2</sup>) |
| --- | --- | --- | --- | --- | --- | --- |
| **Cardiovascular analysis, age ≥65** |  |  |  |  |  |  |
| **All-races** | 930 | 52.8 | 704,808 | 1,335.2 | (reference) | 4,446 |
| <sup>L</sup> NH-White | 925 | 40.4 | 564,536 | 1,398.0 | +5% | 4,444 |
| <sup>L</sup> Hispanic | 191 | 4.1 | 40,086 | 979.7 | -27% | 4,734 |
| <sup>L</sup> NH-Black | 373 | 4.6 | 70,884 | 1,525.6 | +14% | 4,561 |
| **Cancer analysis, age ≥50** |  |  |  |  |  |  |
| **All-races** | 920 | 115.2 | 565,287 | 490.7 | (reference) | 4,444 |
| <sup>L</sup> NH-White | 914 | 82.8 | 441,725 | 533.6 | +9% | 4,442 |
| <sup>L</sup> Hispanic | 198 | 11.5 | 34,621 | 302.0 | -38% | 4,714 |
| <sup>L</sup> NH-Black | 375 | 11.7 | 62,127 | 530.8 | +8% | 4,557 |

**Footnotes:** Values represent the Health Service Areas retained for each analysis after applicable data-availability and suppression criteria. Population and mortality estimates are for 2018-2019. Rates are crude, and race-specific strata are prespecified subsets and do not collectively represent the complete all-races population. HSA, Health Service Area; NH, non-Hispanic; UV, ultraviolet radiation.

### Cardiovascular Mortality for ages ≥ 65 by ambient UV (adjusted for SVI)

All major cardiovascular and individual cardiovascular outcomes were inversely and significantly associated with ambient UV levels (adjusted for SVI) (Figure 3). In the multi-variate regression of UV levels (adjusted for SVI), and major CVD, there was a significant inverse association as assessed with a β UV/IQR of -68.3 (deaths per 100,000) (p<0.0001). Analysis of individual cardiovascular outcomes followed, with a similar inverse or protective signal for heart disease (β UV/IQR = -55.0) and stroke (β UV/IQR = -8.03). The modelled burden assessed by the analysed population/100,000 × β UV/IQR for the major CVD was -36,043, and when assessed by the individual CVD outcomes was -28,967 and -4,042, respectively (Table 2). These protective findings were consistent with a country-level spatial regression analysis of UV and ischaemic heart disease.^27^

**Figure 3.**
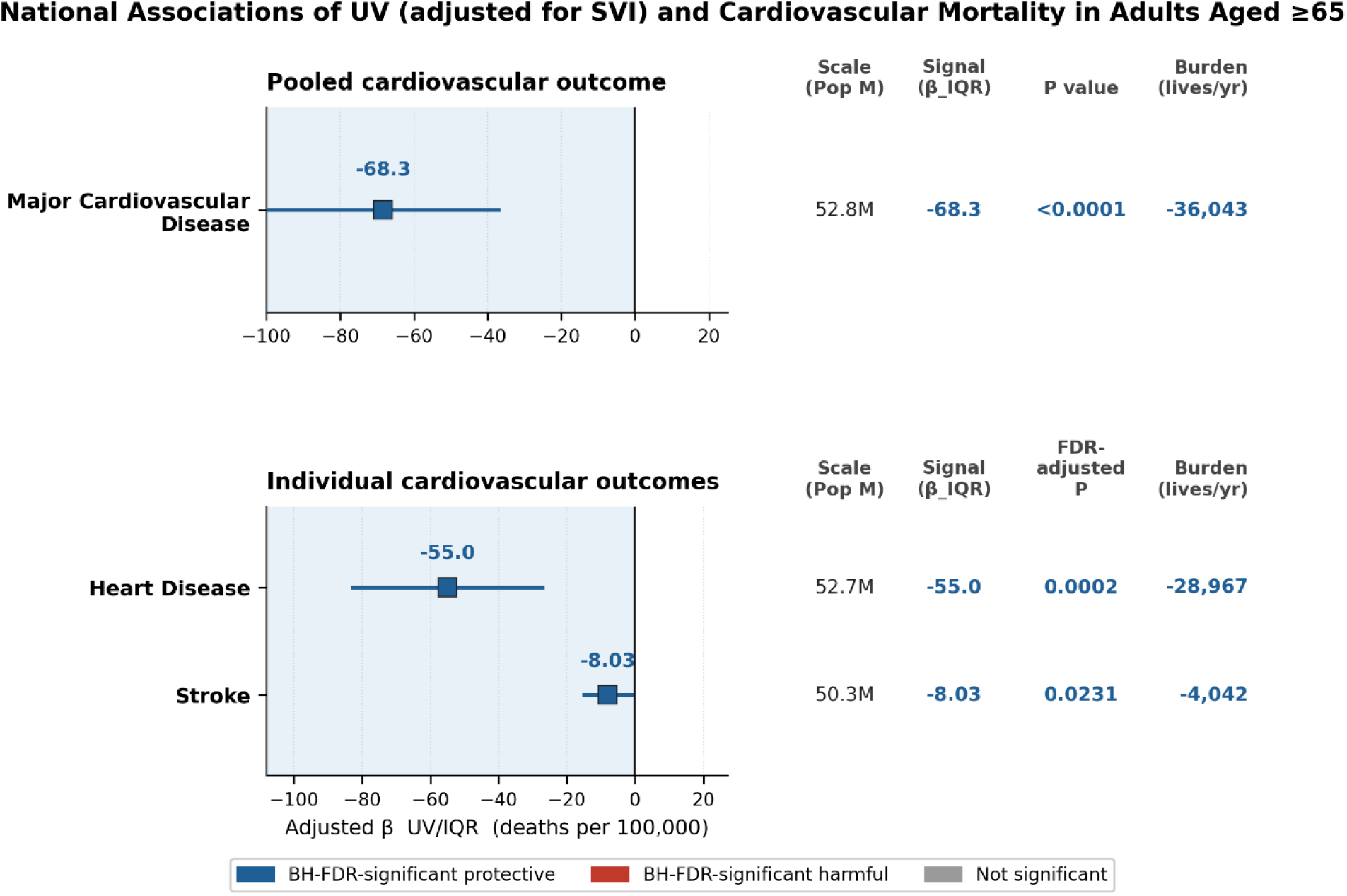
Association between ambient ultraviolet exposure and cardiovascular mortality among US adults aged ≥65 years. Squares represent Social Vulnerability Index-adjusted mortality-rate differences associated with an interquartile-range increase in long-term ambient ultraviolet exposure (β UV/IQR); horizontal lines represent 95% confidence intervals. Negative estimates indicate lower mortality with higher ultraviolet exposure. Scale denotes the analysed population; Signal denotes β UV/IQR; FDR-adjusted P denotes Benjamini–Hochberg false discovery rate (FDR) significant inverse and positive associations; and Burden denotes the modelled annual mortality difference across the ultraviolet interquartile range. Blue indicates statistically significant inverse associations. Estimates use 2018-2019 crude mortality. IQR, interquartile range; SVI, Social Vulnerability Index; UV, ultraviolet.

**Table 2.** Scale, signal, and modelled burden of associations between ambient ultraviolet irradiance and cardiovascular mortality among US adults aged ≥65 years, 2018-2019. Scale is represented by the number of Health Service Areas, annual population, and annual deaths. Signal is the Social Vulnerability Index-adjusted mortality-rate difference associated with an interquartile-range increase in ultraviolet irradiance (β/IQR), expressed as deaths per 100,000 with 95% confidence intervals. Modelled burden was calculated as Σ[(β/IQR × annual population)/100,000]; percentage burden was calculated relative to annual deaths. Individual protective and harmful totals include only component outcomes with FRD-adjusted P < 0.05. Major cardiovascular disease was analysed separately and excluded from the individual-outcome net. Negative values indicate lower mortality associated with higher ultraviolet irradiance. *Asterisk identifies FDR-adjusted P < 0.05 and outcomes included in the modelled burden. CI, confidence interval; HSA, Health Service Area; IQR, interquartile range.Σ[(*β*/IQR × annual population)/100,000] for FDR − adjusted P < 0.05

| Endpoint | Scale of Analysis |  |  | Signal and Significance |  | Modelled Burden* |  |
| --- | --- | --- | --- | --- | --- | --- | --- |
| Outcome | n HSAs | Annual population | Annual deaths | $\beta$/IQR (deaths/100k) (95% CI) | FDR-Adjusted P | Burden (deaths/year) | % of annual deaths |
| Major Cardiovascular Disease | 930 | 52,785,912 | 704,808 | -68.3 (-99.5: -37.1) | <0.001* | -36,043 | -5.1% |
| Individual outcomes (n=2 tests) |  |  |  |  |  |  |  |
| Heart Disease | 924 | 52,710,400 | 525,253 | -55.0 (-82.8: -27.1) | 0.0002* | -28,967 | -5.5% |
| Stroke | 744 | 50,318,454 | 122,455 | -8.03 (-15.0: -1.10) | 0.0462* | -4,042 | -3.3% |
| Individual - Protective |  |  | 647,708 |  |  | -33,010 | -5.1% |
| Individual - Harmful |  |  | 0 |  |  | 0 | 0.0% |
| Net |  |  | 647,708 |  |  | -33,010 | -5.1% |

#### Cardiovascular Disease Mortality for ages ≥ 65 by ambient UV (adjusted for SVI): (stratified by race)

All major cardiovascular and individual cardiovascular outcomes showed divergent signals when deaths were analysed by exclusive races NH-White, Hispanic, and NH-Black, (Figure 4). Major CVD (β UV/IQR = - 74.0) and heart disease (β UV/IQR = -72.6) were inversely and significantly associated with ambient UV only in the NH-Black cohort. Within Stroke, there was a positive and significant correlation ( β UV/IQR =

**Figure 4.**
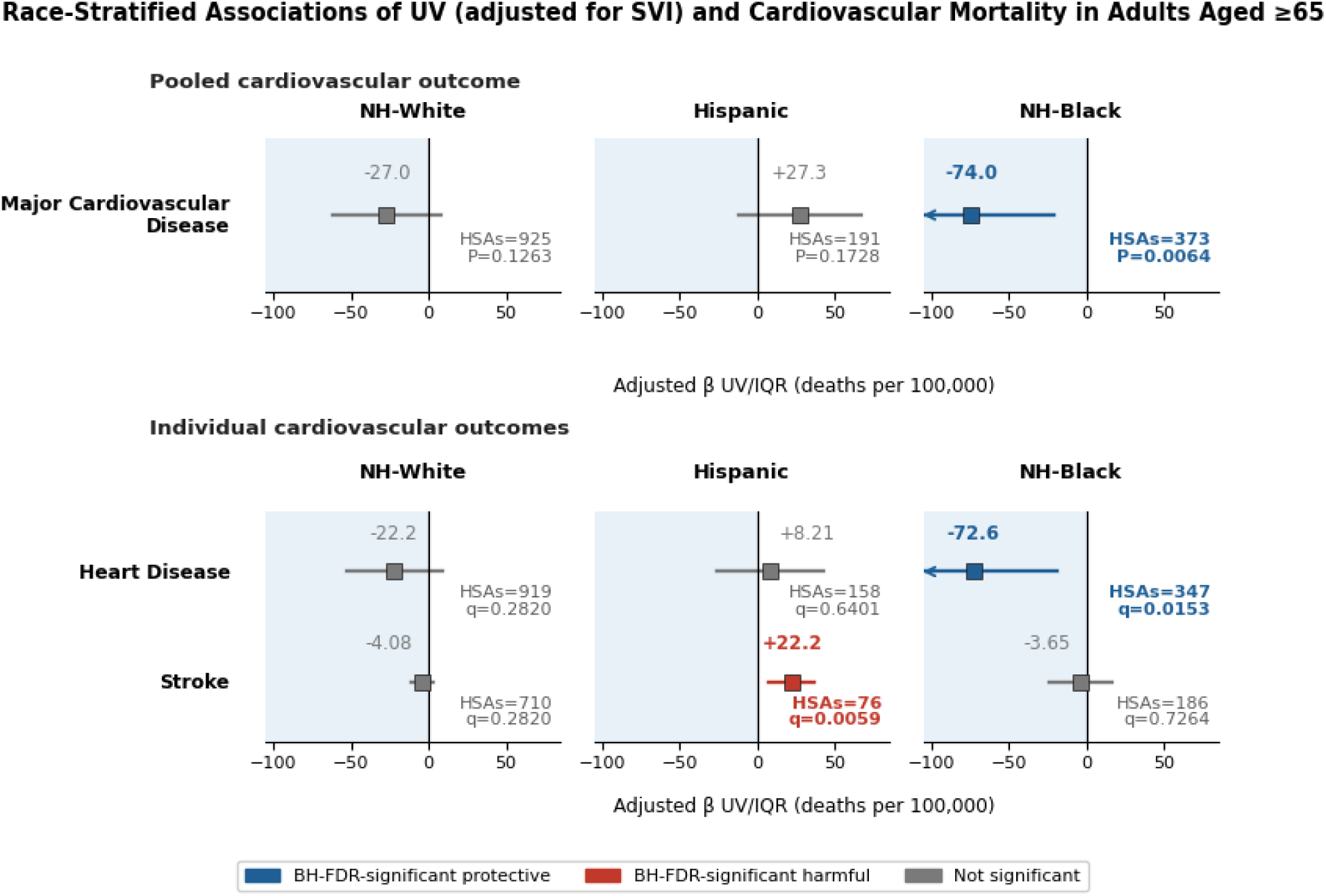
Race-stratified associations between ambient ultraviolet exposure and cardiovascular mortality among adults aged ≥65 years. Squares and horizontal lines show SVI-adjusted mortality-rate differences and 95% confidence intervals per interquartile-range increase in UV. Blue and red indicate Benjamini–Hochberg false discovery rate (FDR) significant inverse and positive associations; grey indicates non-significant associations. Arrows indicate confidence intervals extending beyond the plotted range. Estimates use 2018-2019 crude mortality rates. HSA, Health Service Area; SVI, Social Vulnerability Index; UV, ultraviolet.

+22.2) between Hispanic deaths and UV.

### Cardiovascular Cohort: NH-White Historic Comparison and Corroboration

The outcomes from the previously published UK Biobank differential analyses of the are consistent with our results for both of their reported metrics, lifestyle (active UV - solarium use), and higher ambient UV measures (passive UV - geographic), which reported fully adjusted hazard ratios for cardiovascular mortality associated with UV were 0.77 and 0.81, respectively.^15^

The new fully-adjusted UK Biobank linear UV and cardiovascular mortality shows statistically significant inverse association as assessed by Hazard Ratio 0.80 (0.74–0.87) (p<0.001). Individual CVD outcomes of heart disease of 0.78 (0.71 to 0.86) (p<0.001) and an HR for stroke HR=0.82 of (0.68 to 1.00) (p=0.051) (Table 2).

**Figure 5.**
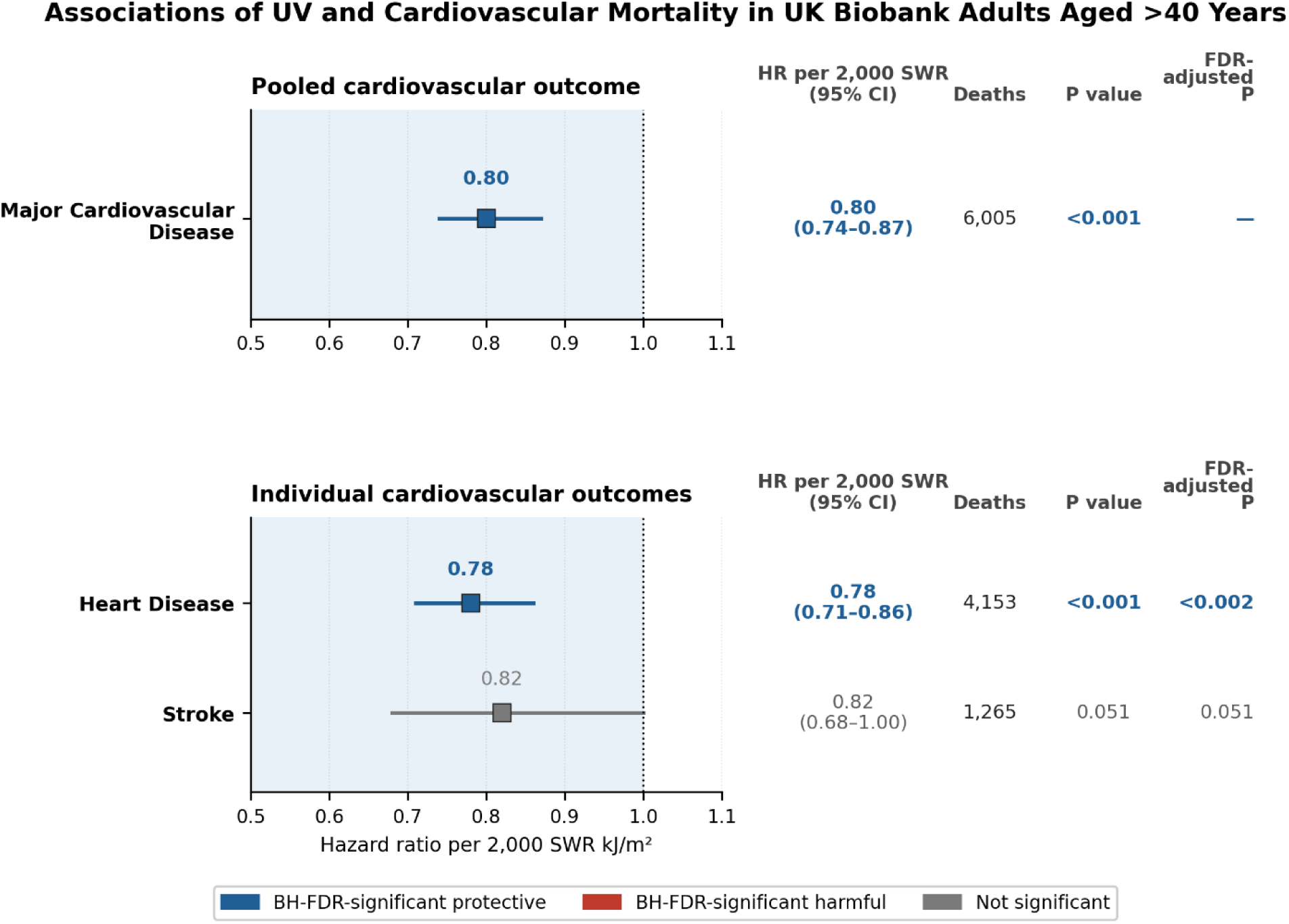
Associations between ambient ultraviolet exposure and cardiovascular mortality in UK Biobank adults aged >40 years. Squares and horizontal lines show fully adjusted hazard ratios (HRs) and 95% confidence intervals per 2,000 kJ/m² increase in annual solar wave radiation (SWR). Blue indicates Benjamini–Hochberg false discovery rate (FDR)-significant inverse associations with mortality; grey indicates non-significant associations. The shaded region denotes HRs <1.0. Models were adjusted for age, sex, Social Vulnerability Index (SVI), and season. SWR, solar wave radiation; SVI, Social Vulnerability Index; UV, ultraviolet.

### Cancer Mortality for ages ≥ 50 by ambient UV levels (adjusted for SVI)

All-cancer mortality and mortality from 9 of 18 individual cancers were inversely and significantly associated with ambient UV levels (adjusted for SVI) (Figure 6). None had a significant and positive correlation. In the multi-variate regression of ambient UV levels (adjusted for SVI) and all-cancer deaths, there was a significant inverse correlation as assessed by UV/IQR with a β UV/IQR of -43.2 (deaths per 100,000) (p<0.0001). Individual cancers followed the same inverse signal, led by lung (-25.8), breast (- 5.32), pancreatic (-4.13) and colon (-3.69); the five further significant outcomes are shown in Figure 6. The modelled burden was -49,789 for all-cancer mortality and -47,940 summed across individual cancers, dominated by lung cancer (-28,791). All outcomes are shown in Figure 6 and Table 3.

**Figure 6.**
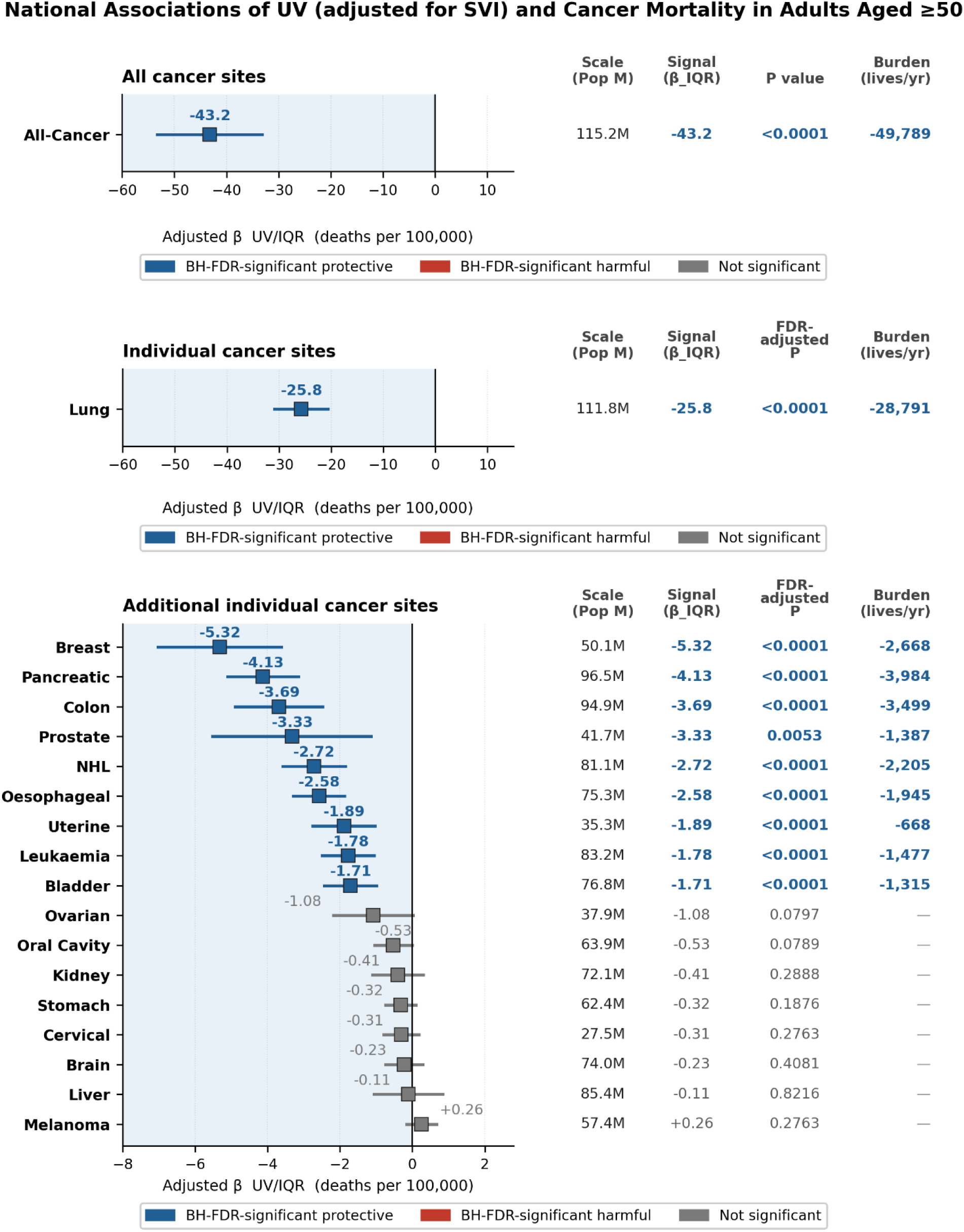
Scale, signal, and modelled burden of the associations between UV and mortality for all-cancer and eighteen individual cancers - Panels A and B are plotted on different scales to show the lung cancer signal (β UV/IQR = -25.8) and the remaining cancers (β UV/IQR < 5). Estimates are SVI-adjusted mortality-rate differences per interquartile-range increase in ambient UV among US adults aged ≥50 years in continental US HSAs, 2018-2019. FDR-adjusted P denotes Benjamini–Hochberg false discovery rate (FDR) significant inverse and positive associations; Blue colour denotes FDR-adjusted P < 0.05. HSA, Health Service Area; IQR, interquartile range; SVI, Social Vulnerability Index; UV, ultraviolet.

**Table 3.**
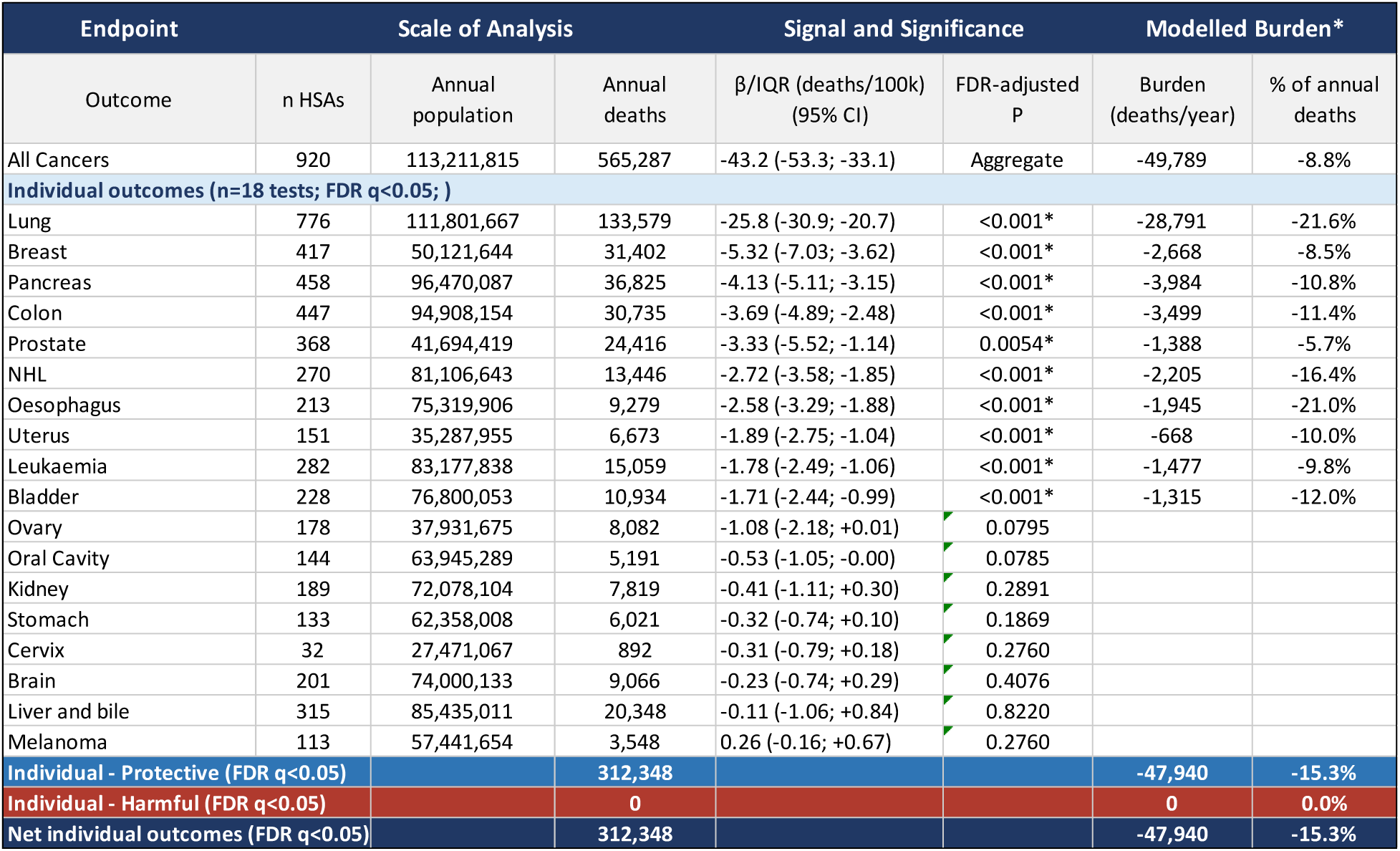
Scale, signal, and modelled burden of associations between ambient ultraviolet irradiance and cancer mortality among US adults aged ≥50 years, 2018-2019. Scale is represented by the number of Health Service Areas, annual population, and annual deaths. Signal is the Social Vulnerability Index-adjusted mortality-rate difference per interquartile range increase in ultraviolet irradiance (β/IQR), with 95% confidence intervals. Definitions of scale, signal, modelled burden and FDR-adjusted P are as for Table 2. Individual protective and harmful totals include only cancer endpoints with FDR-adjusted P < 0.05; the all-cancers aggregate was analysed separately and excluded from the individual net. Negative values indicate fewer deaths associated with higher ultraviolet irradiance. *Asterisk indicates FDR-adjusted P < 0.05 and identifies outcomes included in the modelled burden. CI, confidence interval; HSA, Health Service Area; IQR, interquartile range.FDR − adjusted P < 0.05

| Endpoint |  | Scale of Analysis |  | Signal and Significance |  | Modelled Burden* |  |
| --- | --- | --- | --- | --- | --- | --- | --- |
| Outcome | n HSAs | Annual population | Annual deaths | $\beta$/IQR (deaths/100k) (95% CI) | FDR-adjusted P | Burden (deaths/year) | % of annual deaths |
| All Cancers | 920 | 113,211,815 | 565,287 | -43.2 (-53.3; -33.1) | Aggregate | -49,789 | -8.8% |
| **Individual outcomes (n=18 tests; FDR q<0.05; )** |  |  |  |  |  |  |  |
| Lung | 776 | 111,801,667 | 133,579 | -25.8 (-30.9; -20.7) | <0.001* | -28,791 | -21.6% |
| Breast | 417 | 50,121,644 | 31,402 | -5.32 (-7.03; -3.62) | <0.001* | -2,668 | -8.5% |
| Pancreas | 458 | 96,470,087 | 36,825 | -4.13 (-5.11; -3.15) | <0.001* | -3,984 | -10.8% |
| Colon | 447 | 94,908,154 | 30,735 | -3.69 (-4.89; -2.48) | <0.001* | -3,499 | -11.4% |
| Prostate | 368 | 41,694,419 | 24,416 | -3.33 (-5.52; -1.14) | 0.0054* | -1,388 | -5.7% |
| NHL | 270 | 81,106,643 | 13,446 | -2.72 (-3.58; -1.85) | <0.001* | -2,205 | -16.4% |
| Oesophagus | 213 | 75,319,906 | 9,279 | -2.58 (-3.29; -1.88) | <0.001* | -1,945 | -21.0% |
| Uterus | 151 | 35,287,955 | 6,673 | -1.89 (-2.75; -1.04) | <0.001* | -668 | -10.0% |
| Leukaemia | 282 | 83,177,838 | 15,059 | -1.78 (-2.49; -1.06) | <0.001* | -1,477 | -9.8% |
| Bladder | 228 | 76,800,053 | 10,934 | -1.71 (-2.44; -0.99) | <0.001* | -1,315 | -12.0% |
| Ovary | 178 | 37,931,675 | 8,082 | -1.08 (-2.18; +0.01) | 0.0795 |  |  |
| Oral Cavity | 144 | 63,945,289 | 5,191 | -0.53 (-1.05; -0.00) | 0.0785 |  |  |
| Kidney | 189 | 72,078,104 | 7,819 | -0.41 (-1.11; +0.30) | 0.2891 |  |  |
| Stomach | 133 | 62,358,008 | 6,021 | -0.32 (-0.74; +0.10) | 0.1869 |  |  |
| Cervix | 32 | 27,471,067 | 892 | -0.31 (-0.79; +0.18) | 0.2760 |  |  |
| Brain | 201 | 74,000,133 | 9,066 | -0.23 (-0.74; +0.29) | 0.4076 |  |  |
| Liver and bile | 315 | 85,435,011 | 20,348 | -0.11 (-1.06; +0.84) | 0.8220 |  |  |
| Melanoma | 113 | 57,441,654 | 3,548 | 0.26 (-0.16; +0.67) | 0.2760 |  |  |
| **Individual - Protective (FDR q<0.05)** |  |  | **312,348** |  |  | **-47,940** | **-15.3%** |
| **Individual - Harmful (FDR q<0.05)** |  |  | **0** |  |  | **0** | **0.0%** |
| **Net individual outcomes (FDR q<0.05)** |  |  | **312,348** |  |  | **-47,940** | **-15.3%** |

### Cancer Mortality for ages ≥ 50 by ambient UV levels (adjusted for SVI): Stratified by race

All-cancer and individual cancer outcomes with significant correlations show inverse correlations except one, when deaths were analysed by exclusive races NH-White, Hispanic, and NH-Black (Figure 7). For all-cancer mortality, the NH-White and NH-Black cohorts maintained inverse associations (β UV/IQR = -21.0 and -41.5, respectively). Hispanic all-cancer mortality had a positive (β UV/IQR = +22.7; p=0.0042) but non-significant association with ambient UV levels after Benjamini–Hochberg false discovery rate (FDR) significant inverse (Blue colour) and positive associations (Red colour). Within individual cancers, mortality from lung cancer showed the most significant inverse associations in the NH-White (β UV/IQR = -17.6) and NH-Black (β UV/IQR = -18.2) cohorts between ambient UV levels and mortality. Other significant NH-White associations were inverse (pancreatic, colon, NHL, oesophageal), with melanoma positive as a control (+0.80). Hispanic and NH-Black kidney cancer mortality were positive. Values are given in Figure 7.

**Figure 7.**
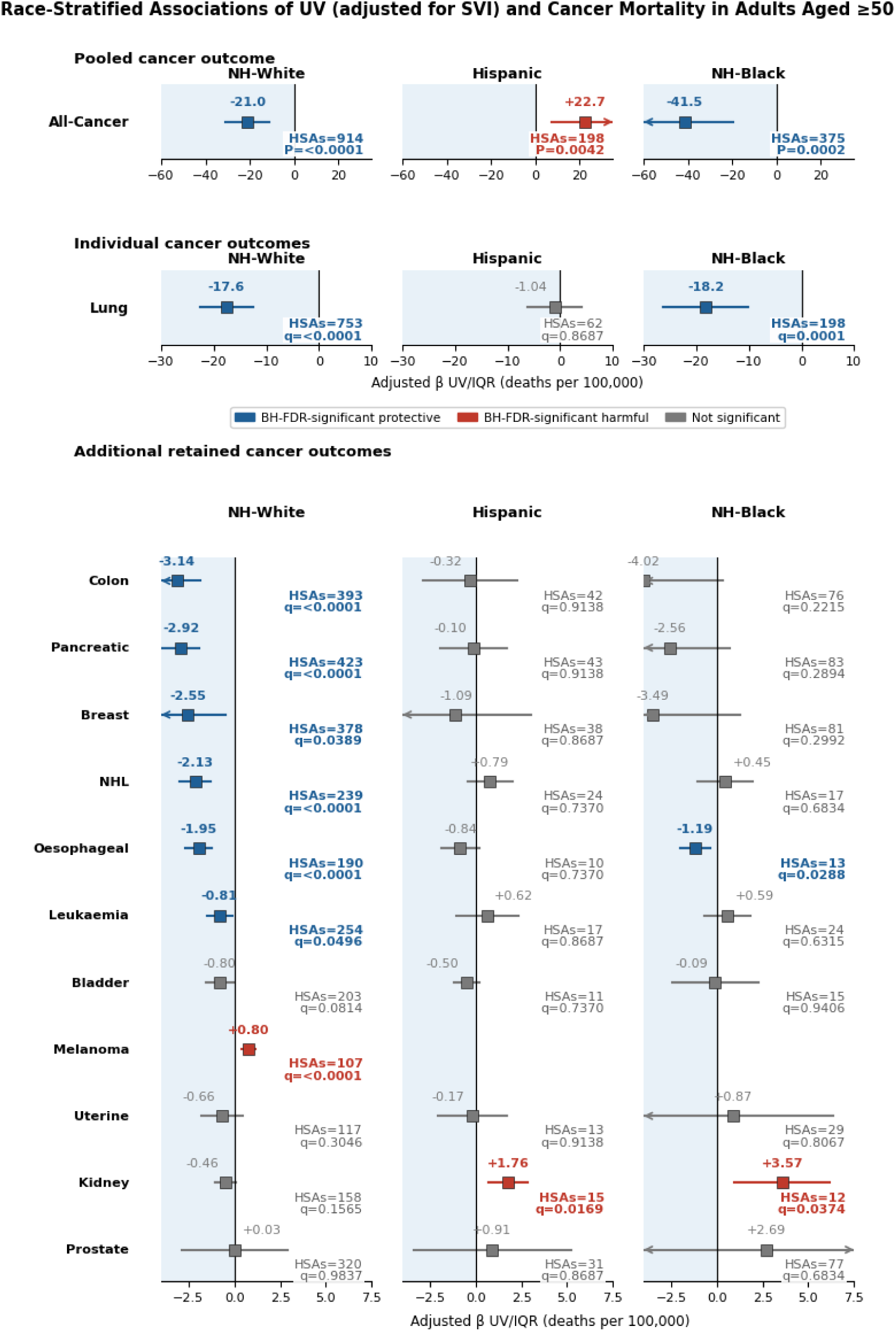
Race-stratified scale and signal of associations between ambient UV and cancer mortality in non-Hispanic White, Hispanic, and non-Hispanic Black populations aged ≥50 years, continental US HSAs, 2018-2019. for cancers significant at the national level, and for those cancers that emerge under race-stratified analysis.

A post-hoc analysis restricting NH-White and NH-Black populations to the same high-UV Hispanic HSAs, and a geographic sensitivity analysis across the 198 HSAs where the Hispanic population resides, have been reported in Supplementary Appendix D (Figure S5). Neither indicated that broad geographic UV distribution alone accounts for the racial and ethnic heterogeneity observed here.

### Cancer Cohort: NH-White Historic Comparison and Corroboration

Within our NH-White cohort and the two additional datasets, we compared UV-mortality associations for all-cancer mortality and all 18 individual body-site-specific cancer outcomes (Figure 8). Because each dataset measured the associations with different methodologies, we used a heatmap to convey direction. All-cancer mortality was concordant, protective, and statistically significant in both the present IQR analysis and the UK Biobank cohort, while Boscoe & Schymura did not report an all-cancer mortality estimate.^2^ Among the individual cancers, twelve showed directional agreement across the two contemporary datasets, ten inverse and two positive, while six were discordant (bladder, colon, leukaemia, breast, brain, and prostate). Leukaemia was discordant because the UV–mortality association was inverse in the historical state analysis and the present NH-White HSA analysis, but directionally positive and non-significant in UK Biobank.

**Figure 8.**
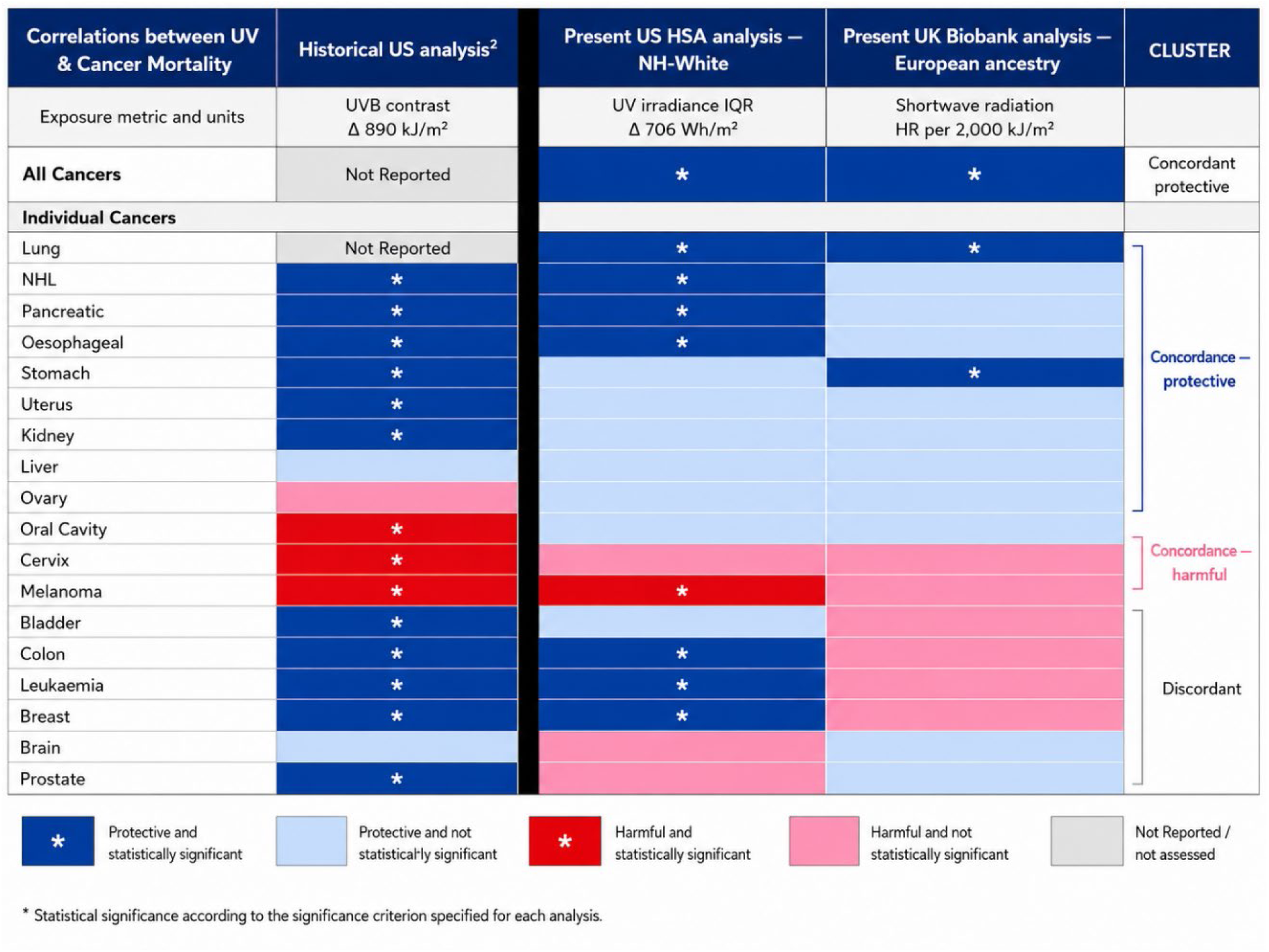
Concordance of ultraviolet and solar exposure associations with cancer mortality in NH-White and European-ancestry populations. Direction and statistical significance are compared across the historical NH-White US state analysis by Boscoe and Schymura (UVB contrast, 890 kJ/m²),² the present NH-White US HSA analysis (UV irradiance IQR, 706 Wh/m²), and the present European-ancestry UK Biobank analysis (HR per 2000 kJ/m² increase in residential shortwave radiation). The pooled all-cancer outcome is shown in the first row. The cluster column classifies each cancer by whether the two contemporary analyses agree in the direction of association; the historical analysis is set apart by the vertical rule and does not enter that classification. Because the exposure metrics, scales, study designs, and effect estimates differ, magnitudes are not directly comparable. Dark blue and red indicate statistically significant protective and harmful associations, respectively; lighter shades indicate corresponding non-significant directions. Asterisks indicate statistical significance after the multiple-testing correction specified for each analysis; the historical analysis did not report multiple-testing-corrected significance, which affects the number of associations marked in that column. Grey indicates that the outcome was not assessed. HR, hazard ratio; HSA, Health Service Area; IQR, interquartile range; NH, non-Hispanic; UV, ultraviolet; UVB, ultraviolet B.

Of the concordant cancers, ten had a protective UV level–mortality association and two (melanoma and cervical cancer) a harmful association. Melanoma reached statistical significance in at least two datasets, while cervical cancer was directionally harmful in all three datasets and statistically significant in one dataset.^28^

Taken together, despite substantial differences in definition of UV levels, outcome modelling, and geographic scale, these datasets provide convergent support for the UV-associated cancer mortality signal. Classifying all 18 individual cancers by the direction of association in the two contemporary datasets gives ten concordant protective, two concordant harmful, and six discordant. The ten concordant protective sites account for 184,747 of the 275,204 annual NH-White cancer deaths represented here (67%) and 8,357 of 13,716 deaths in UK Biobank (61%), against 3,421 and 375 deaths respectively in the two concordant harmful sites. The concordant inverse sites therefore represent most cancer deaths captured in both the NH-White US and UK Biobank comparisons, rather than a collection of rare or isolated outcomes.

The inverse UV–cancer mortality association is not confined to aggregate cancer mortality. Across ecological U.S. analyses and an independent individual-level UK cohort, many cancer-specific associations are directionally concordant despite substantial differences in exposure definition, population, study design and statistical power. Concordance is stronger for direction of association than for formal statistical significance.

### Cardiovascular Disease and Cancer Mortality Burden

Across cardiovascular and cancer mortality outcomes at the national level, these results showed a consistent, strong inverse (protective) association between ambient UV levels and mortality for major CVD, all-cancer, and across the prespecified individual outcomes. Significant positive or harmful associations were identified only in the race-stratified analysis, for Hispanic stroke mortality and NH-White melanoma mortality.

The modelled combined burdens, calculated from the analysed population and the β UV/IQR, were of a substantive magnitude:

- Combined burden by major CVD and all-cancers was -85,832 deaths/year
- Combined burden by individual outcomes of CVD and cancer was -79,563 deaths/year
- Combined burden by race stratified analysis was -49,609 deaths, with Hispanic stroke (+780) and NH-White melanoma (+283) accounting for the only harmful burdens.

## Discussion

### Principal Findings

This study identified consistent associations between long-term ambient UV exposure and reduced cardiovascular and cancer mortality across the United States. The estimated reductions in major cardiovascular disease (CVD) and all-cancer mortality exceeded 5% and 8%, respectively, per interquartile range (IQR) increase in ambient UV levels. The associations were not uniform across individual outcomes, with heart disease dominating in CVD and lung in cancer. Among the 18 individual body-site-specific cancers evaluated, ten demonstrated statistically significant protective associations with ambient UV radiation levels after Benjamini-Hochberg control of the false discovery rate.

The scale of the study enabled race-stratified analyses, in which the NH-White cohort demonstrated the clearest and most numerous protective associations. Melanoma served as the expected positive control and was the only consistently harmful association. Unlike the linear dose-response relationship observed across Fitzpatrick skin types in clinical NB-UVB phototherapy, and the modest gradient of vitamin D production by similar UV exposure,^29^ the population-level associations differed substantially by race and ethnicity.

Among NH-Black populations, the inverse all-cancer association was driven primarily by lung cancer (closely mirroring the magnitude observed among NH-White populations) and secondarily by oesophageal cancer. Kidney cancer was the sole positive cancer association in the NH-Black cohort and was also positive in the Hispanic cohort. This concordance is notable because lung cancer is one of the few malignancies in which the historic mortality disparity between NH-Black and NH-White populations has disappeared over the past three decades.^30^

In UV–mortality analyses, the Hispanic cohort showed a statistically significant positive association for all-cancer mortality (β, +22.7 deaths per 100,000 per IQR; P=0.0042), driven largely by kidney cancer, the only individual cancer to reach significance in this group (β, +1.76; q=0.0169). That reversal of direction against every other stratum is why the Hispanic cohort is reported in its own appendix rather than among the sensitivity analyses: it requires geography, occupational exposure and cancer-site distribution to be treated together before the signal can be interpreted. Those analyses are set out in Supplementary Appendix D4.

### Corroboration

Our findings are consistent with, and extend, previously reported associations between UV exposure, cardiovascular mortality, and cancer mortality. In our two published UK Biobank analyses, both active behavioural UV exposure measured by solarium use and passive environmental exposure measured by residential shortwave radiation were independently associated with lower all-cause, cardiovascular, and cancer mortality.^15,16^ That active sun seeking survival benefits are consistent with the long-term follow-up of lifestyle UV exposure reported in the MISS study.^12–14^ The present manuscript provides an additional level of corroboration by comparing cancer-specific mortality associations across our NH-White U.S. HSA analysis, the newly performed UK Biobank analysis, and the historical U.S. analysis of Boscoe and Schymura.^2^

Concordance was assessed across all 18 individual cancers between the two contemporary datasets, our NH-White US HSA analysis and our UK Biobank analysis, with the historical US state analysis shown alongside as an indication of the level of agreement in the United States two decades earlier. Ten cancers were concordant and protective, two were concordant and harmful (cervix and melanoma), and six (bladder, colon, leukaemia, breast, brain and prostate) were discordant. Concordance here means agreement in the direction of association rather than joint statistical significance. The historical analysis did not apply a correction for multiple testing, which affects the number of associations reaching significance in that column and is the reason it is set apart from the concordance classification.

Across the two contemporary datasets, concordance was stronger for direction of association than for formal statistical significance, as expected given differences in exposure measurement, population structure, outcome counts, and statistical modelling.

### Potential Mechanisms of Action

Our previous works suggest that UV provides a recurring biological stimulus across vascular and immune systems.^9,15^ In this study, we see a significant association between ambient UV and reduced mortality for cancer and cardiovascular disease.

There are multiple potential pathways for connecting ambient UV exposure with the mortality pattern observed here; the supporting experimental, clinical, and mechanistic evidence is detailed in Supplementary Appendix F. The cardiovascular pathway is the most established: cutaneous UV exposure mobilises nitric oxide and produces systemic vascular effects, including vasodilation and lower blood pressure.^48^ A second established pathway is photoimmune regulation, through which UV exposure alters regulatory and effector immune responses locally and systemically.^58,59^ The third pathway, which we hypothesise, concerns cancer immune surveillance: increased mutational burden at the skin^45^ could be driving immune expansion of anti-neoplastic T-cells.

### Clinical Translation

There is a lack of discourse on the risks vs. benefits of UV and mortality. As a modifiable lifestyle exposure, moderate sun-seeking is an actionable option for the population. Decades of sun-avoidance messaging has targeted intermittent burning exposure, but the same message discourages the regular, non-burning exposure on which acclimatisation depends.^31,32^ The skin provides immediate biological feedback through erythema and tanning on its capacity to adapt and produce melanin.

NB-UVB under controlled protocols is a candidate translational therapy beyond dermatology. For cancer, the implications extend beyond prevention because the endpoint in this study was mortality rather than incidence. The observed associations therefore raise the possibility that regular nonburning UV may influence tumour progression, treatment response, recurrence, or survival after diagnosis.

### Limitations of this Study

Ambient UV radiation is a proxy for exposure, and without patient level data, the use of ambient UV has limitations. The modelled confounder of Social Vulnerability Index (SVI) was designed to aggregate vulnerability within communities to prepare for, respond to, and recover from natural or manmade hazards due to the socioeconomic, demographic, and cultural factors that characterise each community. In a recent manuscript of all-cause mortality and SVI across the US with data overlapping our empirical time window, Black-White disparities were not significant in the least vulnerable quartile (AAMR ratio = 1.10, 95% CI: 0.96-1.25, p = 0.16) but remained significant in all other quartiles and for the overall population.^38^

Data from counties were aggregated into Health Service Areas (HSAs). Our intent was to include more counties, as HSAs on average include 2-6 counties, which would increase the number of deaths in an HSA when we applied the SEER suggested low-end cut-off of n=16. We ran the analysis at the county level and confirmed a narrowing of the error bars at the HSA level. Selection of the sub-categories for cardiovascular, heart disease and stroke, captured the dominant reported CVD death classification of heart disease, but limited that discrete number of potentially UV sensitive outcomes. Crude rather than age-adjusted rates were used since age-adjustment rates required breaking the national population averages by age group and race stratum at the county and then HSA levels which substantially limited the number of data points through our filter process. Details are provided in Supplementary Table S5. National annual mortality burden and crude mortality rates by race and ethnicity across the eligible HSAs are provided in Supplementary Table S6; both are descriptive context only.^33^

Selection of the sub-categories of cancer was based on the SEER published cancers of interest (n=21), two of which are childhood cancers, which were excluded by age, and thyroid cancer crude rates were not sufficient to survive initial filters. This limited data set also provides an arbitrary specific cancer subset for analysis. This may have limited our ability to identify discrete UV responsive, and non-UV responsive cancers.

Lastly, UV exposure is highly correlated with other wavelengths of sun exposure, severely diluting any hypotheses generated solely from this work associated with the mechanism of mortality reduction.

### Strengths of this Study

All inputs to the regression were within public datasets within the Centers for Disease Control and Prevention’s Wide-ranging Online Data for Epidemiologic Research (WONDER) and Surveillance, Epidemiology, and End Results (SEER) databases. Aggregating the data in population-weighted HSAs, improved the signal to noise. Use of an Ordinary Least Squares regression model over a Weighted Least Squares model reduced potential double counting. Sensitivity analysis of county vs. HSA, and OLS vs. WLS, univariate analysis of UV vs. mortality, and UV adjusted for SVI and mortality were also performed.

The UK Biobank cohort is a dynamic resource that, unlike the frozen snapshot in time of the US HAS data, can provide continuous updates of patient-level data for future insights.

## Conclusion

Our findings suggest opportunities for intervention at both the lifestyle and therapeutic levels across a discrete set of cardiovascular conditions and cancers. Consistent with long-term Swedish cohort and UK Biobank findings, regular nonburning sun exposure may be associated with net survival benefits compared with strict sun avoidance. In parallel, controlled NB-UVB provides an existing translational platform to test whether photobiologic stimulation can improve cardiovascular and cancer survival. Reconciling the epidemiological and clinical data therefore necessitates challenging the assumption that all UV exposure is biologically equivalent or uniformly harmful; in fact, multiple studies including this one provide evidence suggesting that for some populations, a significant survival benefit would be conferred through moderately increasing sun exposure. The concordance between these population-level associations and outcomes observed under prescribed narrowband UVB, where exposure is independent of lifestyle choice, points to a shared photobiological mechanism and makes controlled phototherapy a practical platform for testing it prospectively.

## Data Availability

US data used in this study are derived from publicly available sources, including CDC WONDER, the National Cancer Institute GIS/AVGLO ultraviolet exposure data, the CDC/ATSDR Social Vulnerability Index, and the SEER/NCHS county-to-Health Service Area crosswalk. UK Biobank participant-level data are available to approved researchers through UK Biobank and were accessed for this study under Application Number 82526. The v29 WONDER Mortality Engine pipeline is available from the corresponding author on reasonable request.

## DATA AVAILABILITY

US data are derived from public sources (NCI GIS/AVGLO; CDC/ATSDR SVI; NCHS/SEER county-HSA crosswalk; CDC WONDER). UK Biobank data sourced from research conducted using the UK Biobank Resource under Application Number 82526.

## Article statements

## Acknowledgements

We thank Craig Ullman for careful proofreading of the manuscript. We also acknowledge the patients and families affected by cancer, including NHL and pancreatic cancer, whose experiences motivated our interest in understanding potentially modifiable environmental influences on cancer mortality.

## Contributors

JM conceived and designed the study and drafted the manuscript. CE conducted parallel statistical analysis and proposed the cancer vaccine hypothesis, PH and BW interpreted the findings and critically revised the manuscript. RW, CD, and AS acquired and curated the UK Biobank data and its analysis. All authors approved the final manuscript and accepted accountability for the work. JM is the guarantor.

## Ethical Approval

Analyses of US public-use datasets (CDC WONDER and SEER) were deemed exempt from institutional review board review as they use fully de-identified, aggregate, publicly available data and do not constitute human subjects research under 45 CFR 46. Analyses of UK Biobank data were conducted under Application 82526. UK Biobank has ethical approval from the Northwest Multi-centre Research Ethics Committee (REC reference 11/NW/0382), and all participants provided written informed consent at enrolment.

## Data Sharing Statement

The v29 WONDER Mortality Engine pipeline is available from the corresponding author on reasonable request.

## Dissemination to participants and related patient and public communities

The US analysis used de-identified, aggregate, publicly available data with no enrolled participants. The UK Biobank analysis used data from consented participants under Application Number 82526, and results will be returned to UK Biobank in accordance with its access terms. Findings will be disseminated to scientific, clinical and patient communities through peer-reviewed publication and open-access summaries.

## Use of artificial intelligence

The authors used a large language model (Anthropic Claude) to assist with manuscript preparation, specifically verification of references against CrossRef and PubMed, repair of citation and cross-reference integrity, and editing for length and clarity. No artificial intelligence tool was used to generate or interpret study data, to perform statistical analysis, or draft scientific conclusions. The authors reviewed and edited all content and take responsibility for its accuracy and integrity.

## Transparenc

We affirm that the manuscript is an honest, accurate, and transparent account of the study being reported; that no important aspects of the study have been omitted; and that any discrepancies from the study as planned (and, if relevant, registered) have been explained.

## Provenance and peer review

Not commissioned; not yet peer reviewed (preprint).

## Online Supplementary Appendix: Methods, Tables, and Figures

**For manuscript: Cardiovascular and cancer mortality inversely associated with long-term ambient ultraviolet exposure across US Health Service Areas with UK Biobank corroboration**

### Supplementary Appendix A. Vitamin D supplementation and mortality evidence

This appendix summarises randomised trials in which cholecalciferol was compared with placebo, drawn from two structurally different evidence streams. The first comprises general-population prevention trials in unselected or community-dwelling adults, which randomised tens of thousands of participants and followed incident disease or death over several years. The second comprises smaller trials in patients already diagnosed with cancers of the types represented in the study cohort, in which vitamin D was added to standard oncological care and outcomes were expressed as relapse-free, event-free, or treatment-free survival.

These two streams differ fundamentally in their endpoints, which limits direct comparison between them. Mortality was rarely the prespecified primary endpoint: among the trials listed here, only D-Health prespecified all-cause mortality, while the remaining prevention trials were designed around incident cancer, cardiovascular events, or fractures, and the disease-specific trials around relapse or disease progression. Cause-specific mortality estimates, where they exist, generally come from secondary or long-term follow-up analyses. Effect estimates are reproduced as published, are not pooled, and are accompanied by the endpoint status for each trial.

None of the randomised placebo-controlled trials summarised in Supplementary Table S1 demonstrated a statistically significant benefit for a prespecified primary cardiovascular, cancer, mortality, or survival endpoint. Taken with the observational associations reported in the main manuscript, this pattern suggests that vitamin D status may be a marker of parallel benefit from ambient ultraviolet exposure rather than the explanation for the observed UV-mortality associations.

**Supplementary Table S1.**
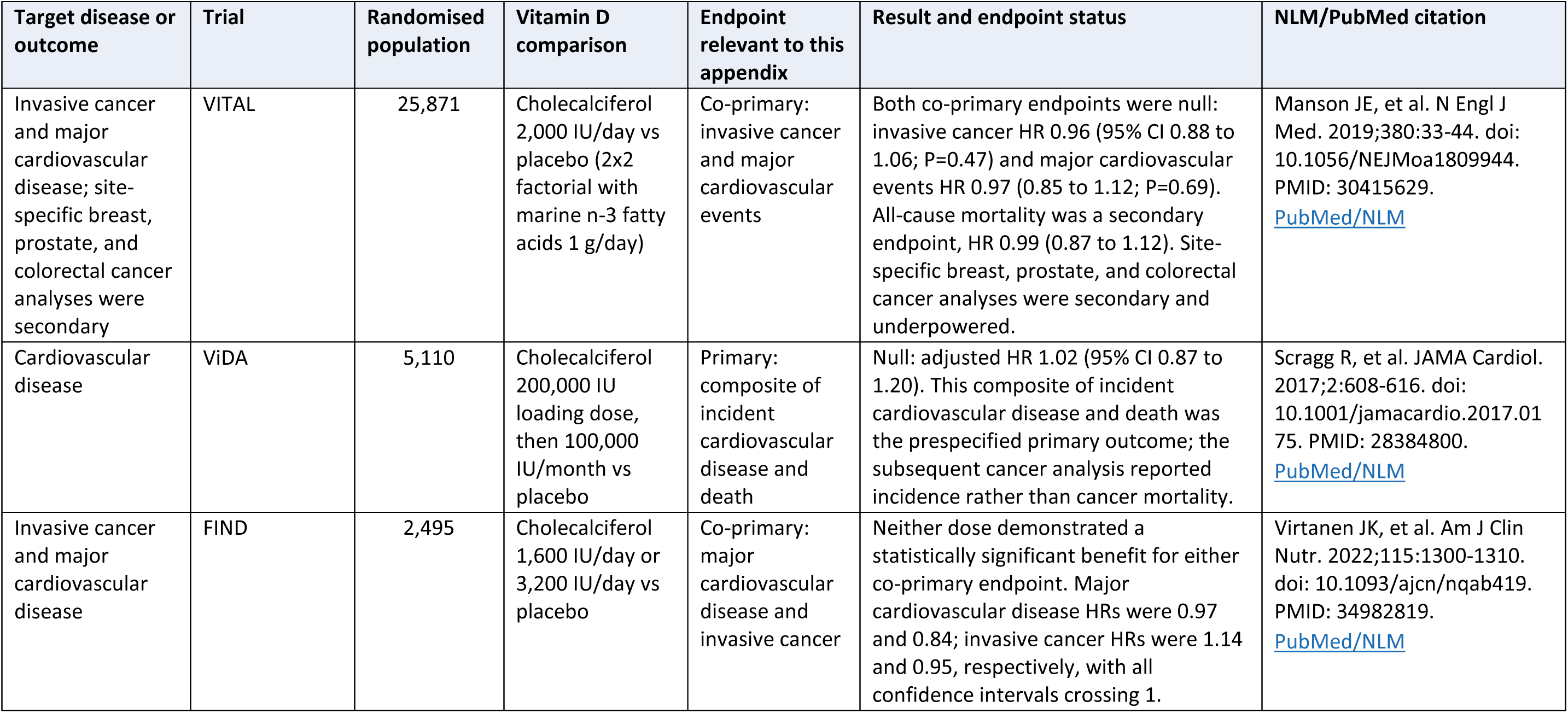
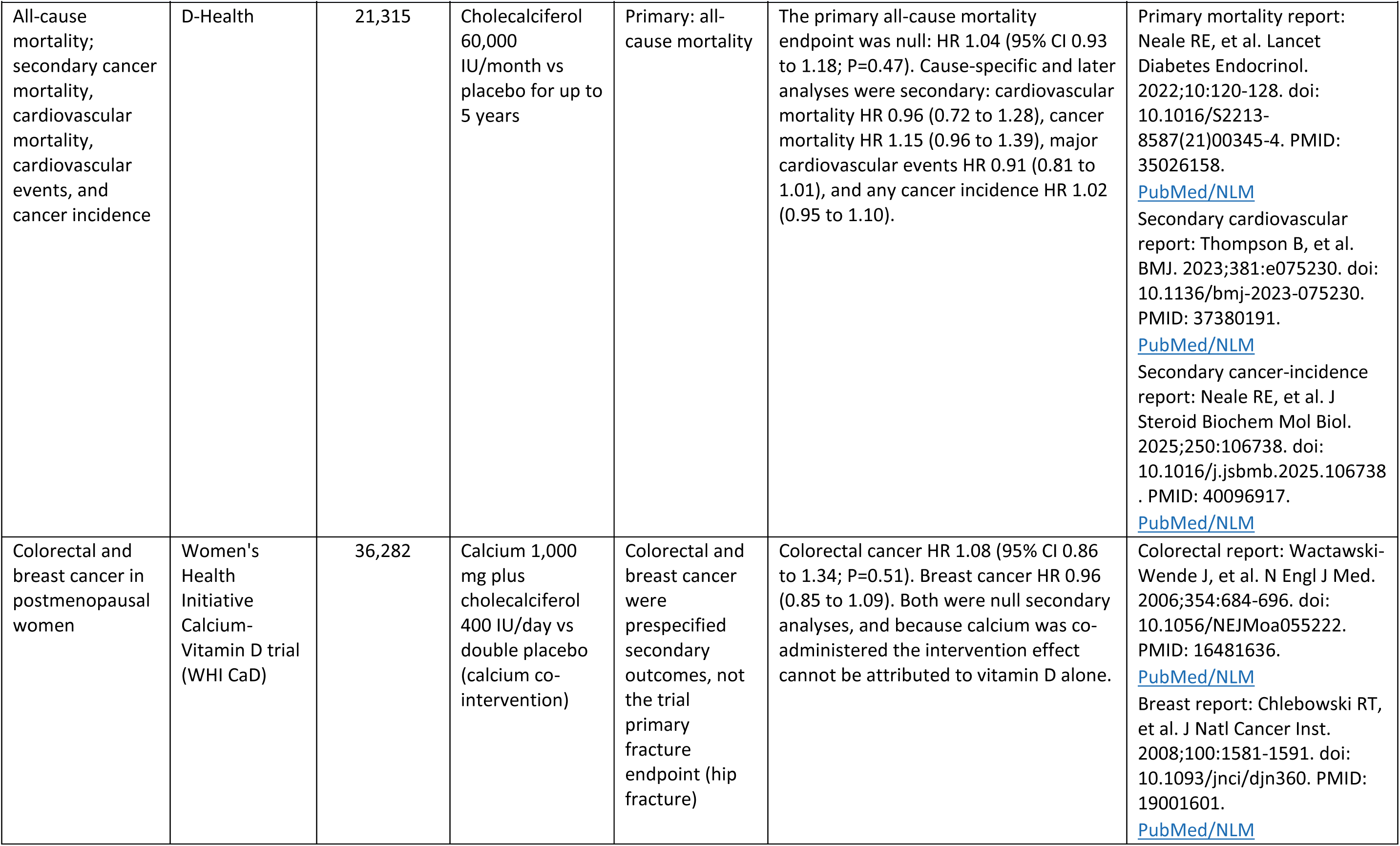
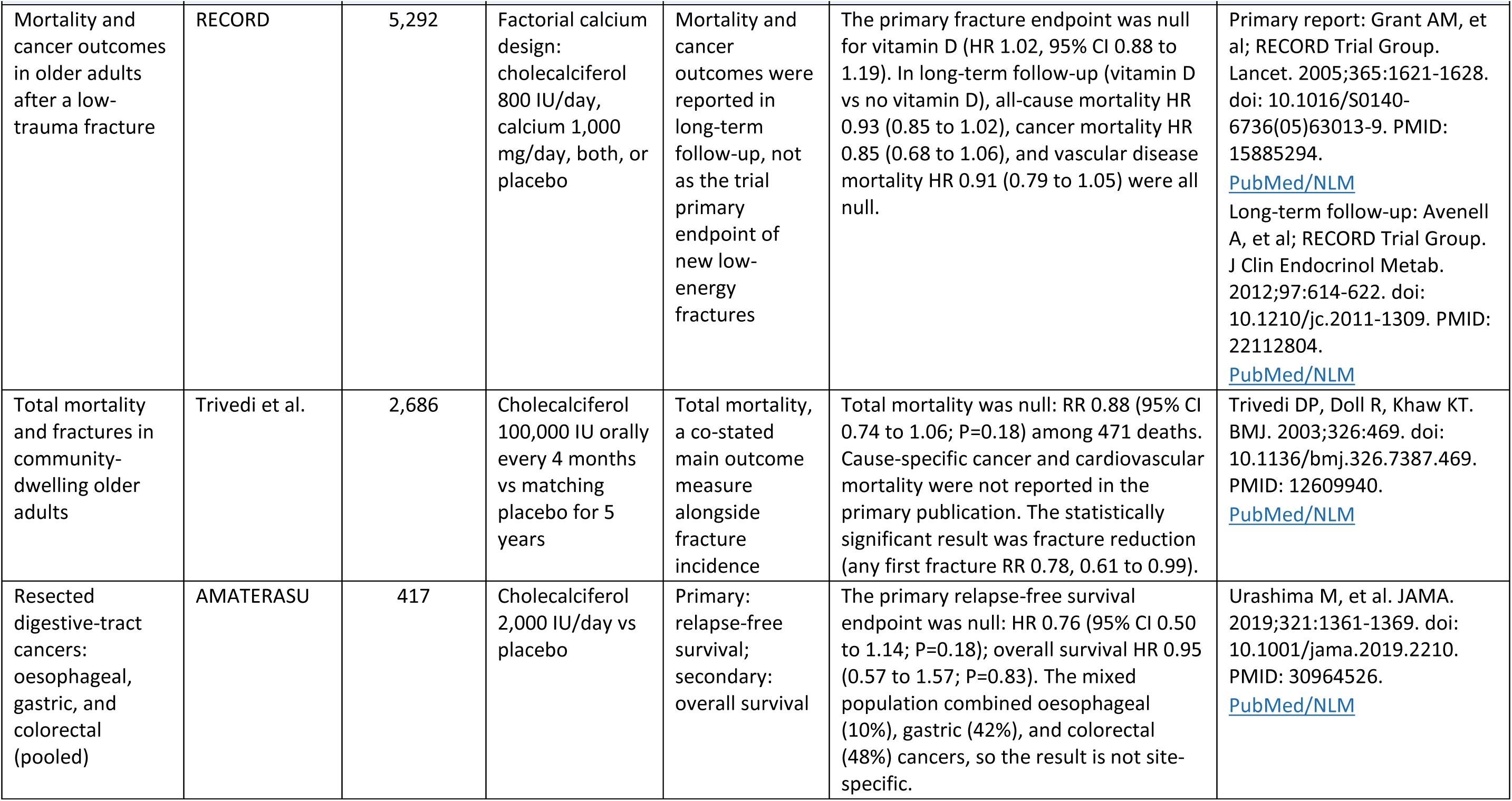
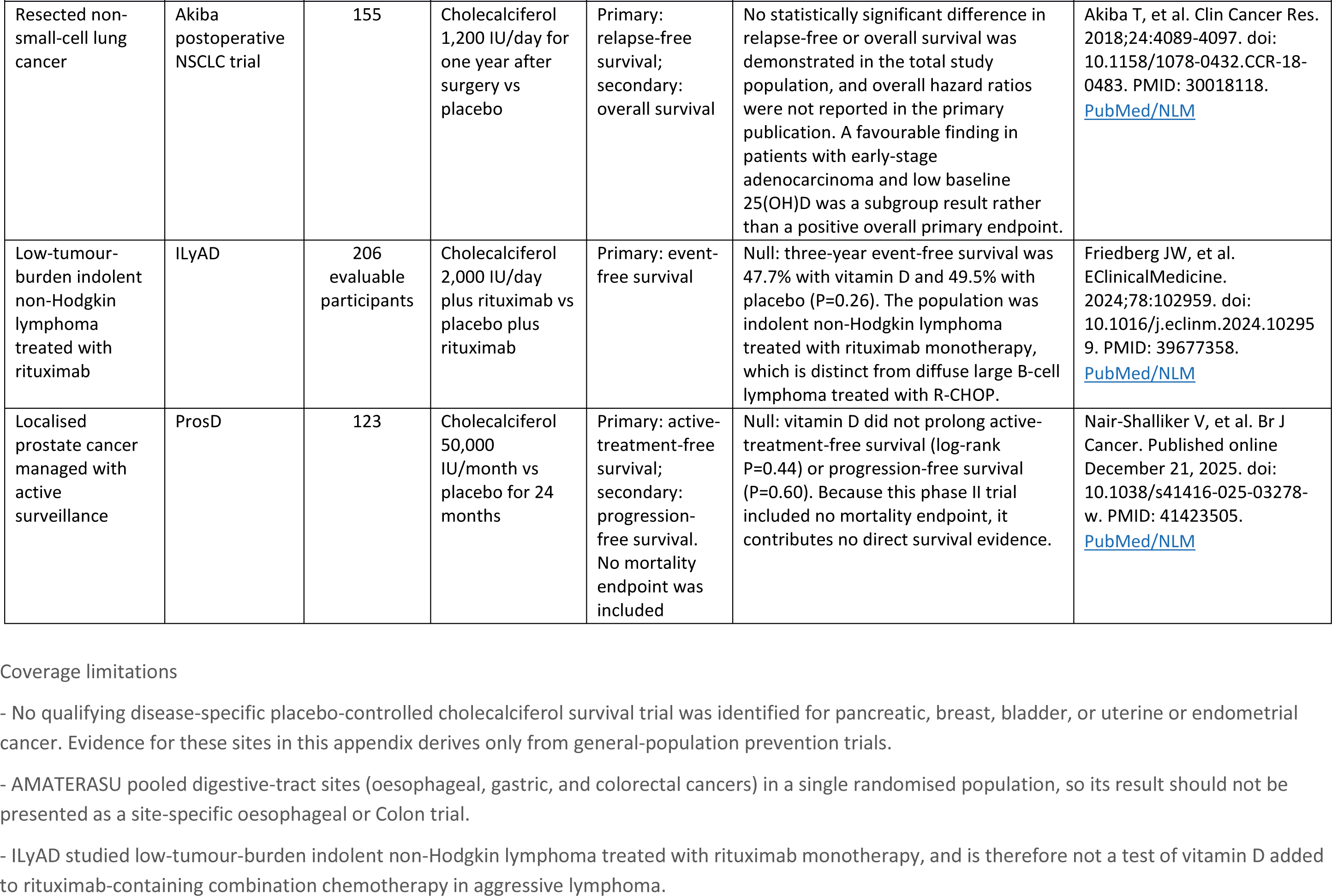
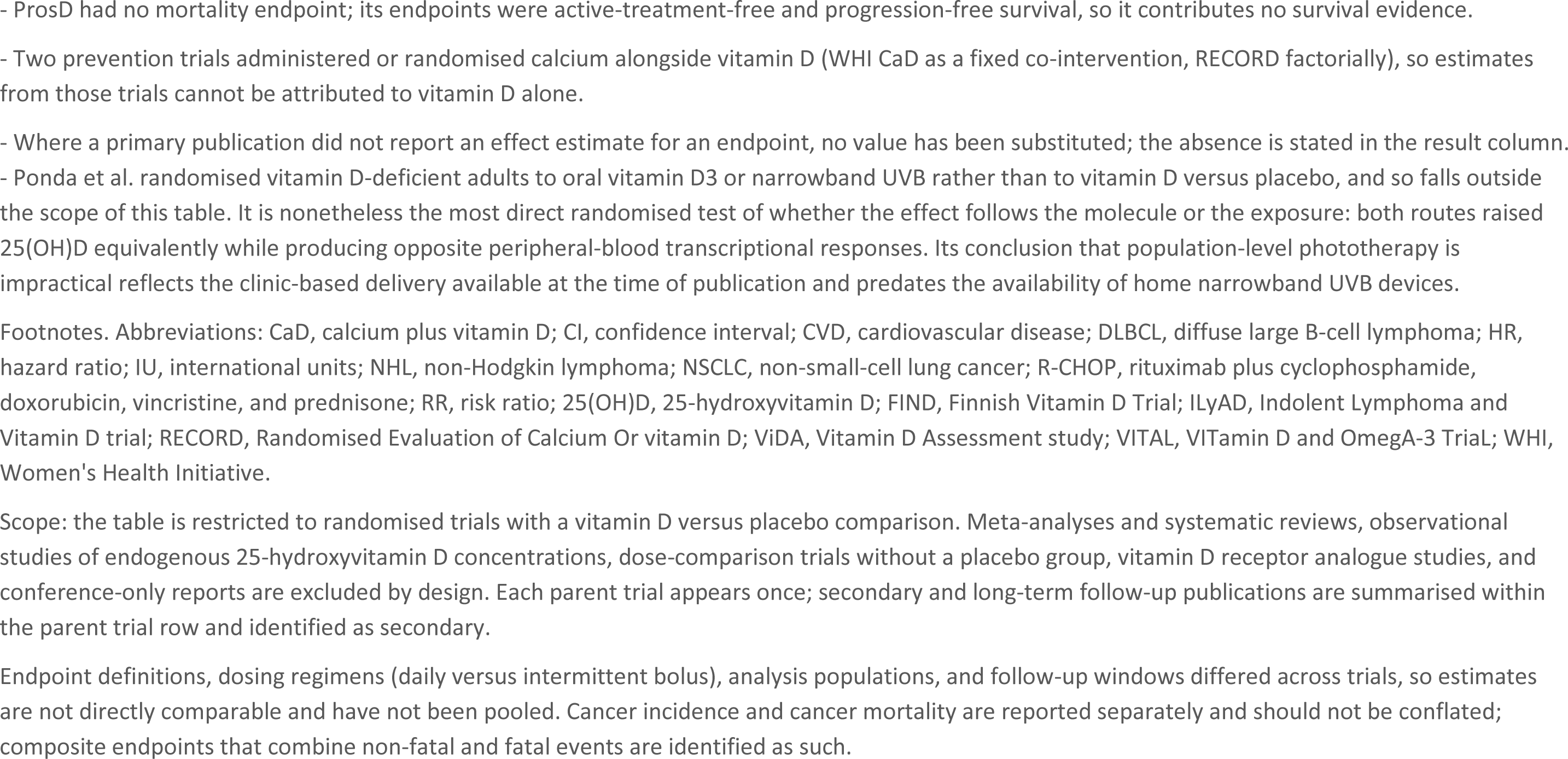
Randomised placebo-controlled vitamin D trials relevant to cardiovascular and cancer outcomes examined in the study.

### Supplementary Appendix B. US and UK mortality endpoint definitions

Mortality endpoints analysed in the US Health Service Area analysis were defined by underlying cause-of-death codes of the International Classification of Diseases, 10th Revision (ICD-10), as recorded in CDC WONDER. Supplementary Table S2 lists the aggregate and component endpoints and the verified code sets applied to each.

CDC WONDER suppresses county-year cells containing 1-9 deaths.^34^ Released counts were summed across constituent counties and 2018-2019, allowing cumulative observed HSA counts to exceed the 16-death stability threshold. Suppressed county-year counts remained unavailable and could not be recovered through aggregation.

County UV and SVI values were population-weighted to the HSA level. Available deaths and corresponding populations were summed across constituent counties and years, and crude mortality rates were calculated from these totals. HSA-outcome cells with fewer than 16 observed deaths were excluded according to the SEER/US Cancer Statistics stability criterion.^24^ Analyses without this threshold were examined as sensitivity analyses (Supplementary Methods). The primary cardiovascular and cancer datasets included 930 and 920 HSAs, representing 52.8 million adults aged ≥65 years and 115.2 million adults aged ≥50 years, respectively.

WONDER mortality and population were downloaded by all-races, and by exclusive race/ethnicity strata NH-White, Hispanic, and NH-Black. Hispanic includes respondents of any race who self-identified as Hispanic or Latino; NH-White and NH-Black are restricted to respondents who did not. Each stratum has its own county-level death counts, its own age-eligible population denominator, and therefore its own HSA-level crude rate, eligible HSA set, and β UV/IQR. The strata are not adjustments to a pooled model; they are four independent regressions run on four independent slices of the data to isolate the UV-mortality signal within each race group. The same β UV/IQR and burden calculations were repeated for each outcome (using their own β UV/IQR and eligible HSA set) and for each race stratum (all-races, NH-White, NH-Black, Hispanic).

**Supplementary Table S2.** Mortality endpoints and corresponding International Classification of Diseases, 10th Revision codes used to identify underlying causes of death in CDC WONDER, United States, 2018-2019.

| Underlying cause-of-death category | Common Usage / Abbreviation | ICD-10 code(s) |
| --- | --- | --- |
| **Aggregate cardiovascular endpoint** |  |  |
| Diseases of the circulatory system | Major Cardiovascular disease (CVD) | I00-I99 |
| **Individual cardiovascular endpoints** |  |  |
| Diseases of heart | Heart disease | I00-I09, I11, I13, I20-I51 |
| Cerebrovascular diseases | Stroke | I60-I69 |
| **Aggregate cancer endpoint** |  |  |
| Malignant neoplasms | All-cancer | C00-C97 |
| **Individual cancer endpoints** |  |  |
| Trachea, bronchus and lung | Lung | C33-C34 |
| Colon | Colon | C18 |
| Pancreas | Pancreas | C25 |
| Breast | Breast | C50 |
| Prostate | Prostate | C61 |
| Non-Hodgkin lymphoma | NHL | C82-C85 |
| Oesophagus | Oesophagus / Oesophageal | C15 |
| Bladder | Bladder | C67 |
| Corpus uteri and uterine, part unspecified | Uterine | C54-C55 |
| Melanoma of skin | Melanoma | C43 |
| Kidney and renal pelvis | Kidney | C64-C65 |
| Brain | Brain | C71 |
| Liver and intrahepatic bile ducts | Liver | C22 |
| Stomach | Stomach | C16 |
| Lip, oral cavity and pharynx | Oral cavity | C00-C14 |
| Ovary | Ovary / Ovarian | C56 |
| Leukaemia | Leukaemia | C91-C95 |
| Cervix uteri | Cervix / Cervical | C53 |

Footnotes: ICD-10, International Classification of Diseases, 10th Revision; CVD, cardiovascular disease; NHL, non-Hodgkin lymphoma. Codes were applied to the underlying cause-of-death field. Cardiovascular outcomes were evaluated among adults aged 65 years or older and cancer outcomes among adults aged 50 years or older. Diseases of the circulatory system and all cancers represent aggregate endpoints; the corresponding individual diseases and cancer sites are component endpoints. ICD-10 definitions were verified against the query criteria recorded in each CDC WONDER source-file footer. CDC WONDER data were retrieved 3-6 June 2026.

**Supplementary Figure S1.**
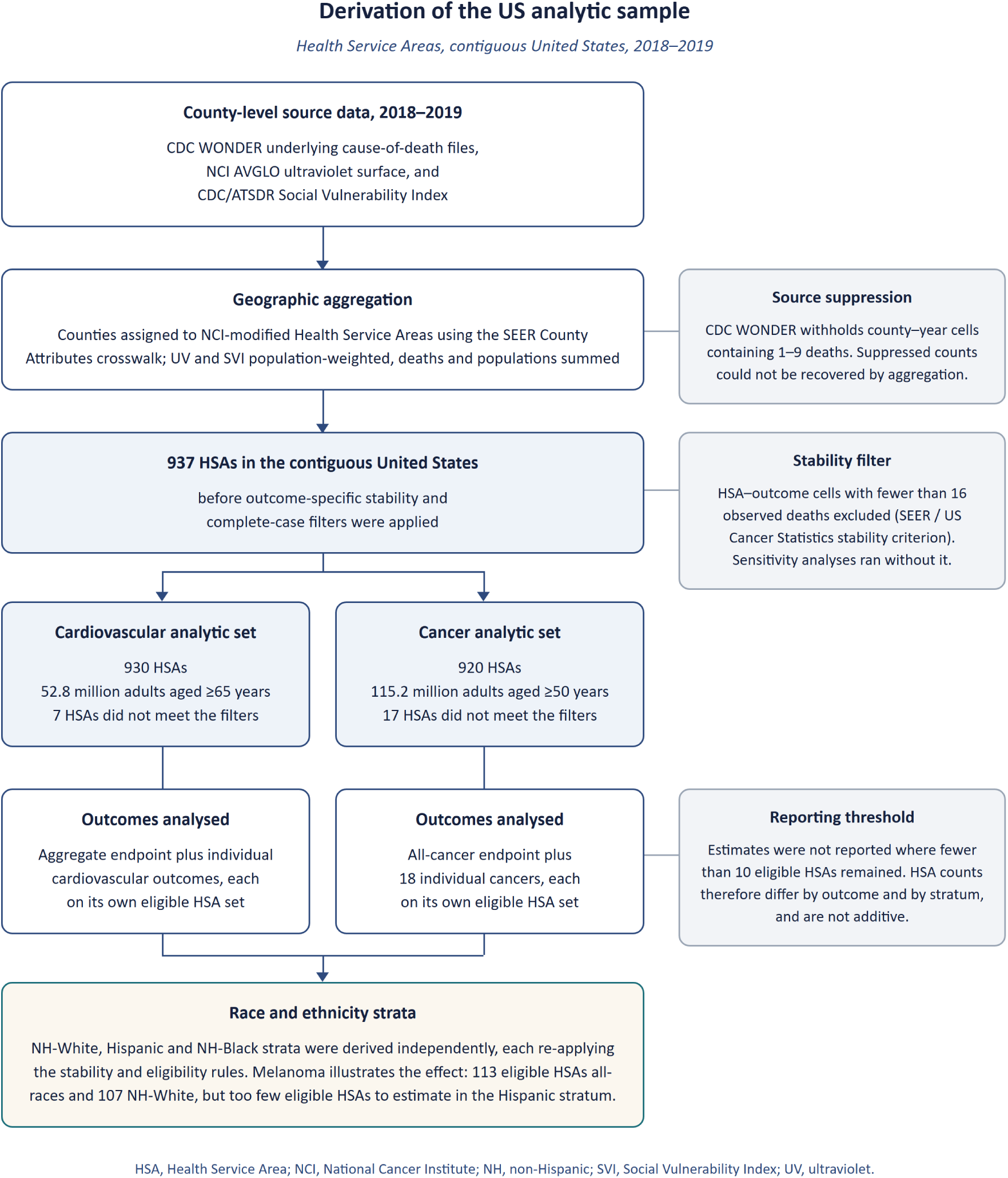
Derivation of the US analytic sample. Health Service Areas (HSAs) in the contiguous United States, 2018-2019. Counts are outcome-specific: HSA-outcome cells with fewer than 16 observed deaths were excluded under the SEER / US Cancer Statistics stability criterion, and estimates were not reported where fewer than 10 eligible HSAs remained. HSA counts therefore differ by outcome and by stratum and are not additive. Race and ethnicity strata were derived independently, each re-applying the stability and eligibility rules.

**Supplementary Figure S2.**
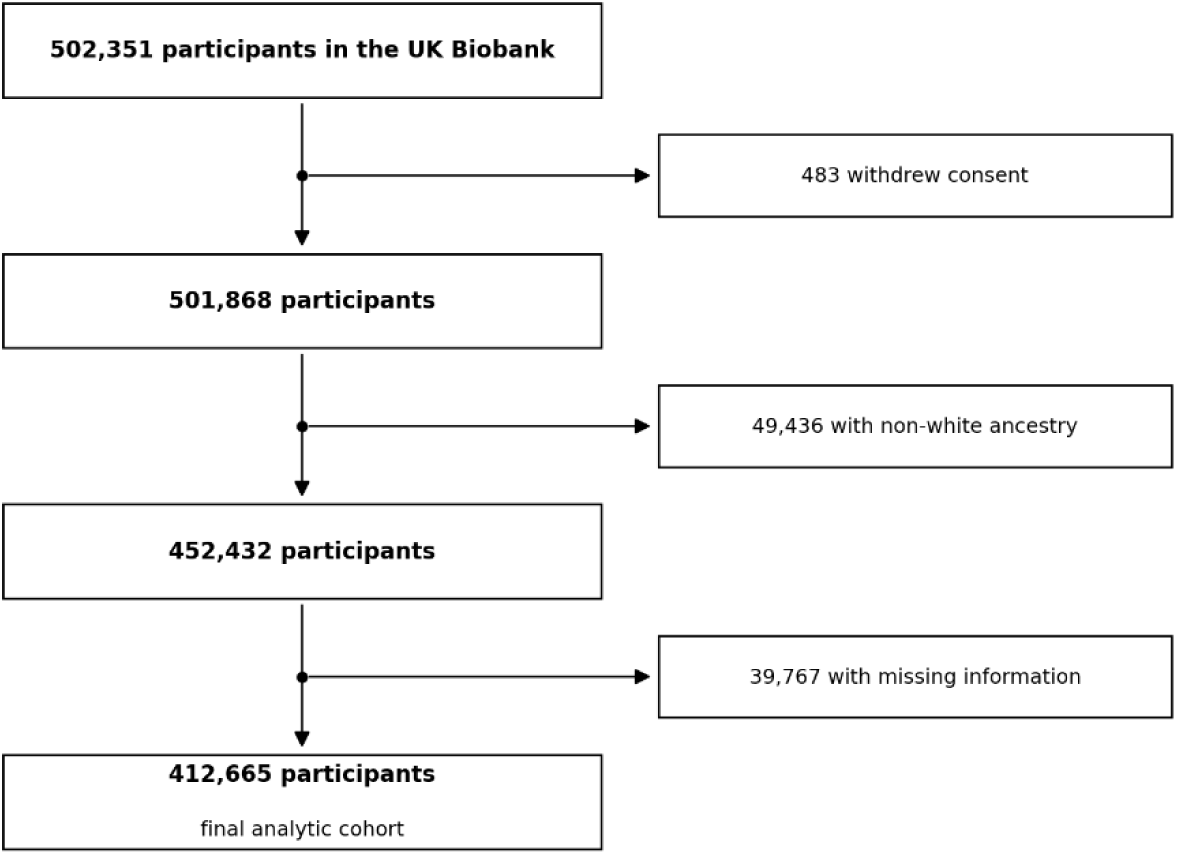
Flowchart of UK Biobank participant information.

### Supplementary Appendix C. US sensitivity and validation analyses

#### C1. Reciprocal ultraviolet and social vulnerability adjustment

Ambient ultraviolet (UV) exposure and the Social Vulnerability Index (SVI) are spatially correlated across US Health Service Areas, so each can act as a confounder of the other. Two model families were therefore compared for every endpoint. Univariate models regressed the HSA-level annual crude mortality rate on one exposure only, either ambient UV exposure or SVI. Mutually adjusted models (traditional multivariable models) included both exposures simultaneously, so that the coefficient for each exposure is estimated with the other held constant. All models were fitted by ordinary least squares at HSA level using 2018-2019 mortality data, with HC1 heteroskedasticity-robust inference, and coefficients are expressed as the difference in annual crude mortality rate per 100,000 person-years associated with an interquartile-range increase in the focal exposure.

The 2018 CDC/ATSDR Social Vulnerability Index (SVI) served as the area-level covariate for socioeconomic and demographic deprivation, available for download at the county level from the CDC/ATSDR SVI interactive portal,^19^ combining 15 American Community Survey indicators across four themes (socioeconomic status, household composition, minority status and language, and housing and transportation) into a single percentile rank per county. Recent work by Ganatra and colleagues has shown graded increases in county-level mortality from cancer, cardiovascular disease, and comorbid cancer– cardiovascular disease across SVI quartiles, supporting SVI as a cardio-oncology relevant measure of structural vulnerability.^35^ Although SVI was developed for emergency preparedness, it has since been empirically validated as a predictor of population mortality across both endpoint families addressed in this analysis. For cardiovascular disease, Khan and colleagues linked county-level SVI to CDC WONDER mortality across all 3,143 U.S. counties and reported an 84% higher rate of premature cardiovascular death in the highest SVI quartile relative to the lowest, with a dose–response across ischemic heart disease, stroke, hypertension, and heart failure.^36^ For cancer, a 2023 systematic review of 31 SVI studies, 15 of which examined mortality, established SVI as a reliable area-level instrument across the cancer care continuum, with higher SVI consistently associated with worse mortality in breast, lung, and colorectal cancer.^37,38^

Benjamini–Hochberg false discovery rate (FDR) correction was applied independently by tier. The top categories (major cardiovascular disease and all-cancer) were tested at α = 0.05, while the individual-outcome tier (all-race, α = 0.05/number of tests) and the race-stratified tier (α = 0.05/number of tests) used a dynamic denominator equal to the number of conditions in that parent passing both the ≥16-event stability rule and the ≥10-eligible-HSA gate, which avoids over-correcting when WONDER suppresses rare endpoints at the source.

Raw β^ s the slope of the regression for deaths per 100,000 per 1 Wh/m² UV. To rescale β^ to a more relevant environmental measure, the interquartile range (IQR) was applied (β_IQR = β^ × IQR), where the IQR is the gap on the x-axis of the regression between the 25th- and 75th-percentile of annual UV. Across the 930 continental US HSAs modelled, the UV IQR is 706 Wh/m² (Q25 = 4,062, median = 4,394, Q75 = 4,769) and the SVI IQR is 0.3916 (See Figure 2).

For the burden calculation of modelled reduced or increased mortality within the filtered dataset, after individual β UV/IQRs were calculated for each outcome, they are multiplied by their unique analysed population and divided by 100,000, so the modelled burden only addresses the population from which it was derived. Net burden represents the sum across all eligible HSAs within each family of tests.

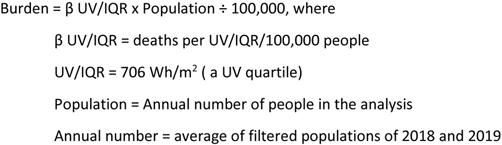

To provide context, an approximate one IQR, 706 Wh/m² UV, is close to the difference in annual UV between the HSA centred on Boston, MA (4,002 Wh/m²) and the one centred on Atlanta, GA (4,681 Wh/m²).

**Supplementary Table S3.** Single-exposure and mutually adjusted (traditional univariate and multivariable) associations of ambient ultraviolet exposure and social vulnerability with cardiovascular and cancer mortality, all races, US Health Service Areas, 2018-2019.

| Mortality endpoint | HSAs (n) | Ambient UV exposure: change in crude rate per IQR increase |  | Social vulnerability: change in crude rate per IQR increase |  |
| --- | --- | --- | --- | --- | --- |
|  |  | Single-exposure beta (p value) | Mutually adjusted beta (p value) | Single-exposure beta (p value) | Mutually adjusted beta (p value) |
| Cardiovascular endpoints, adults aged 65 years or older |  |  |  |  |  |
| Major cardiovascular disease (aggregate) | 930 | -12.9 (0.336) | -68.3* (1.8 × 10<sup>-5</sup>) | 82.5* (3.7 × 10<sup>-7</sup>) | 119.9* (5.0 × 10<sup>-10</sup>) |
| Heart disease | 924 | -8.8 (0.461) | -55.0* (1.1 × 10<sup>-4</sup>) | 69.3* (8.9 × 10<sup>-7</sup>) | 99.4* (3.1 × 10<sup>-9</sup>) |
| Stroke | 744 | 4.7 (0.156) | -8.0* (0.023) | 22.9* (7.5 × 10<sup>-8</sup>) | 27.9* (2.4 × 10<sup>-9</sup>) |
| Cancer endpoints, adults aged 50 years or older (Benjamini–Hochberg FDR, q < 0.05) |  |  |  |  |  |
| All cancers (aggregate) | 920 | -13.5* (0.002) | -43.2* (4.6 × 10<sup>-17</sup>) | 39.0* (9.7 × 10<sup>-14</sup>) | 63.3* (1.6 × 10<sup>-24</sup>) |
| Lung | 776 | -7.3* (8.6 × 10<sup>-4</sup>) | -25.8* (3.9 × 10<sup>-23</sup>) | 24.2* (8.6 × 10<sup>-20</sup>) | 39.8* (3.9 × 10<sup>-41</sup>) |
| Pancreas | 458 | -2.6* (4.1 × 10<sup>-9</sup>) | -4.1* (1.0 × 10<sup>-16</sup>) | 1.2 (0.108) | 4.0* (7.2 × 10<sup>-7</sup>) |
| Breast | 417 | -2.0 (0.019) | -5.3* (9.9 × 10<sup>-10</sup>) | 5.8* (8.6 × 10<sup>-5</sup>) | 9.3* (1.5 × 10<sup>-9</sup>) |
| Colon | 447 | -0.4 (0.506) | -3.7* (1.9 × 10<sup>-9</sup>) | 6.9* (2.0 × 10<sup>-16</sup>) | 9.2* (3.8 × 10<sup>-24</sup>) |
| Prostate | 368 | -0.5 (0.642) | -3.3 (0.003) | 5.7* (1.1 × 10<sup>-5</sup>) | 8.0* (7.3 × 10<sup>-8</sup>) |
| Liver | 315 | 2.8* (1.5 × 10<sup>-8</sup>) | -0.1 (0.822) | 8.2* (2.7 × 10<sup>-18</sup>) | 8.3* (3.0 × 10<sup>-15</sup>) |
| Leukaemia | 282 | -1.3* (5.4 × 10<sup>-5</sup>) | -1.8* (1.2 × 10<sup>-6</sup>) | 0.3 (0.687) | 1.5 (0.030) |
| NHL | 270 | -1.7* (6.1 × 10<sup>-7</sup>) | -2.7* (7.3 × 10<sup>-10</sup>) | 1.1 (0.051) | 3.2* (2.5 × 10<sup>-6</sup>) |
| Bladder | 228 | -1.2* (4.8 × 10<sup>-4</sup>) | -1.7* (3.6 × 10<sup>-6</sup>) | 0.5 (0.373) | 1.8 (0.003) |
| Oesophagus | 213 | -2.3* (2.2 × 10<sup>-11</sup>) | -2.6* (8.4 × 10<sup>-13</sup>) | -0.9 (0.186) | 1.1 (0.122) |
| Brain | 201 | -0.3 (0.151) | -0.2 (0.385) | -0.5 (0.204) | -0.3 (0.496) |
| Ovary | 178 | -0.6 (0.182) | -1.1 (0.053) | 0.9 (0.245) | 1.7 (0.064) |
| Kidney | 189 | 0.4 (0.162) | -0.4 (0.257) | 2.4* (7.4 × 10<sup>-6</sup>) | 2.7* (9.3 × 10<sup>-6</sup>) |
| Uterine | 151 | -1.6* (6.8 × 10<sup>-6</sup>) | -1.9* (1.5 × 10<sup>-5</sup>) | -0.6 (0.420) | 1.0 (0.259) |
| Stomach | 133 | 0.4 (0.059) | -0.3 (0.135) | 2.2* (2.1 × 10<sup>-8</sup>) | 2.5* (2.3 × 10<sup>-9</sup>) |
| Oral cavity | 144 | -0.3 (0.212) | -0.5 (0.048) | 0.4 (0.369) | 0.9 (0.071) |
| Melanoma | 113 | -0.0 (0.955) | 0.3 (0.230) | -0.7 (0.046) | -0.9 (0.026) |
| Cervix | 32 | -0.1 (0.665) | -0.3 (0.218) | 0.3 (0.347) | 0.7 (0.143) |

HSA, Health Service Area; IQR, interquartile range; NHL, non-Hodgkin lymphoma; SVI, Social Vulnerability Index; UV, ultraviolet. All rows are all-races (overall) estimates; no race-stratified rows are shown. Single-exposure models included one exposure only; mutually adjusted models included ambient UV exposure and SVI simultaneously. Betas are differences in annual crude mortality rate per 100,000 person-years per interquartile-range increase in the focal exposure (UV IQR 705.96 units; SVI IQR 0.3916). An asterisk denotes statistical significance at the FDR-adjusted threshold specified for the endpoint family. Values are reproduced from the approved model outputs that generated Supplementary Figure S3; none were recalculated. Confidence limits are not reported here because the approved all-races outputs and the corresponding figure report point estimates, standard-error-based p values, and significance flags only.

**Supplementary Figure S3.**
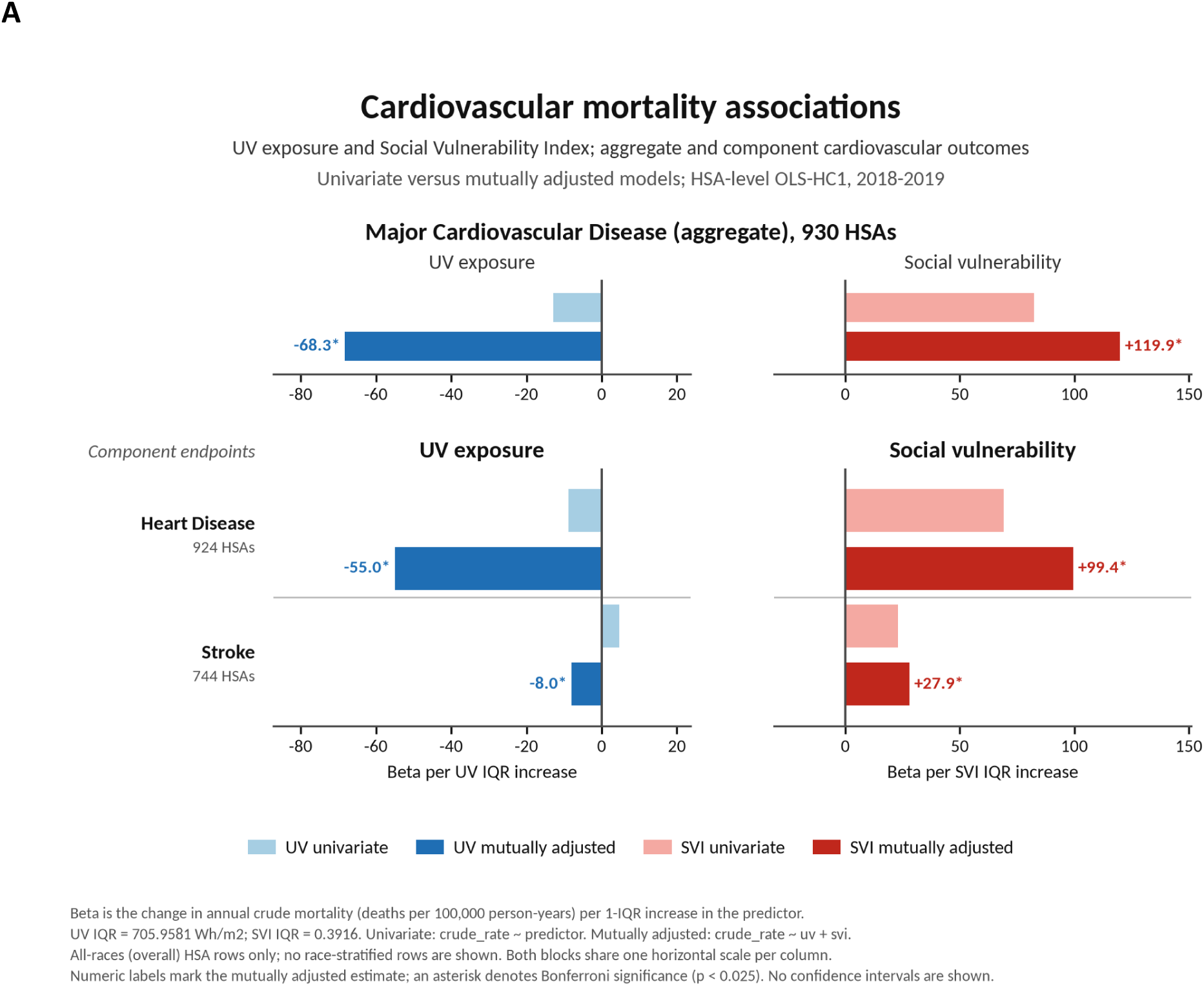
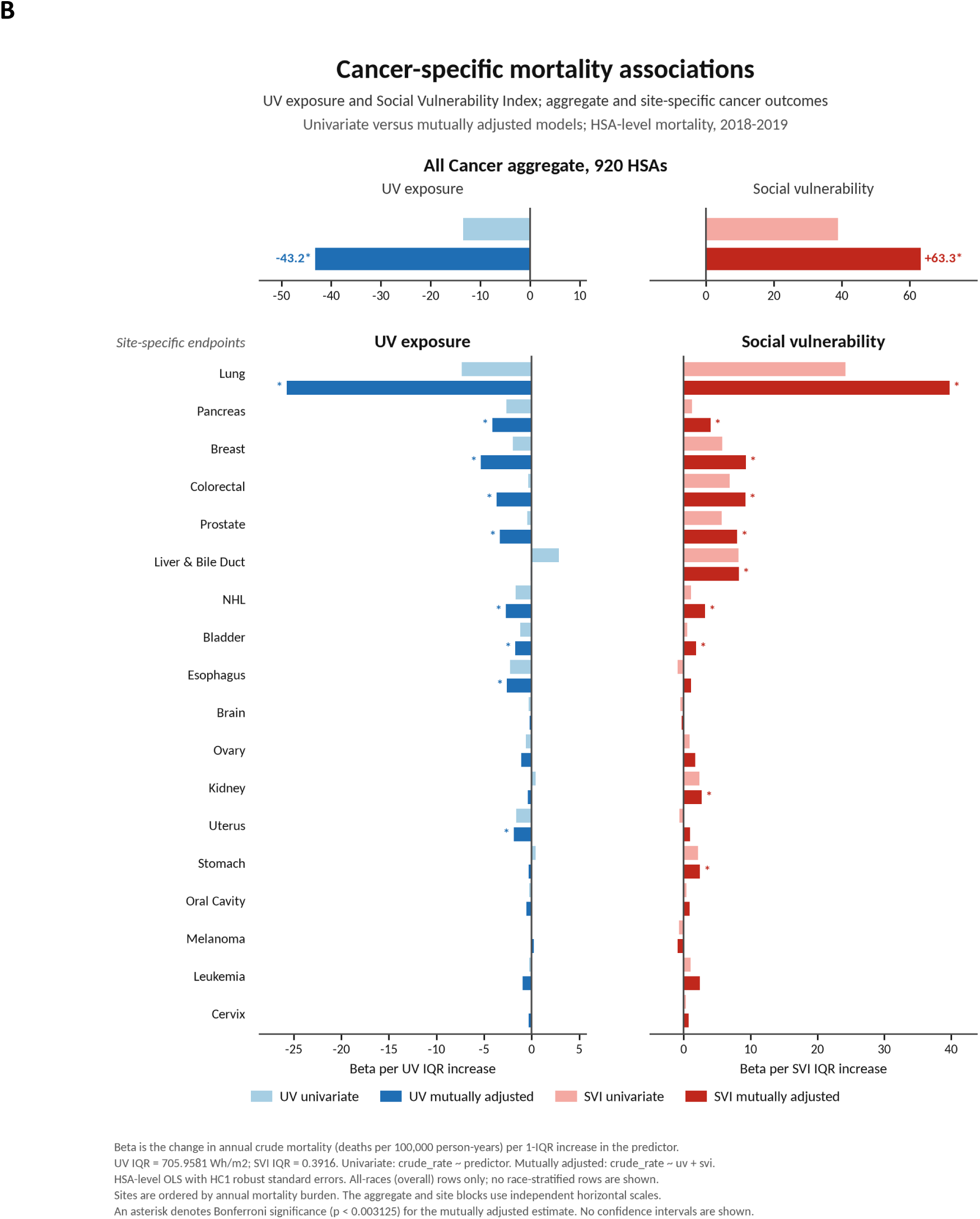
Single-exposure (univariate) and mutually adjusted coefficients of association, Beta per UV IQR, for ambient ultraviolet exposure and social vulnerability with cardiovascular and cancer mortality. Panel A presents the aggregate major cardiovascular disease endpoint and the component heart disease and stroke endpoints. Panel B presents the aggregate all-cancer endpoint and site-specific cancer endpoints. Both panels use HSA-level mortality data for 2018-2019. Light bars show single-exposure (univariate) estimates and dark bars show estimates mutually adjusted for the other exposure. Estimates are differences in annual crude mortality rates per 100,000 person-years associated with an interquartile-range increase in the focal exposure. Numeric labels identify mutually adjusted estimates for the cardiovascular endpoints. Asterisks denote FDR-adjusted significance using the thresholds specified within each panel. Confidence intervals are not displayed. HSA, Health Service Area; IQR, interquartile range; NHL, non-Hodgkin lymphoma; SVI, Social Vulnerability Index; UV, ultraviolet.

#### C2. Linear versus curvilinear fitting sensitivity

##### Supplementary methods

Because ecological UV-mortality models commonly assume a linear relationship, quadratic and restricted cubic spline models were used to assess alternative functional forms.

Primary HSA-level models estimated SVI-adjusted associations between annual ambient UV exposure and crude mortality rates using OLS with HC1 heteroskedasticity-robust standard errors. Models were fitted overall and for NH-White, NH-Black, and Hispanic populations.

Quadratic models added a centred squared UV term. Interior turning points were calculated as *UV*^∗^ = −*β*_1_/(2*β*_2_)when supported by the fitted model.

##### Supplementary results

Cancer associations were predominantly linear and are not detailed further. Cardiovascular models showed greater curvature, primarily reflecting a steeper inverse slope in the highest UV quartile rather than attenuation at higher exposure or evidence of an optimal threshold. Linear models were retained for consistent estimation and the prespecified burden calculations. Representative cardiovascular fits are shown in Supplementary Figure S4.

**Supplementary Figure S4.**
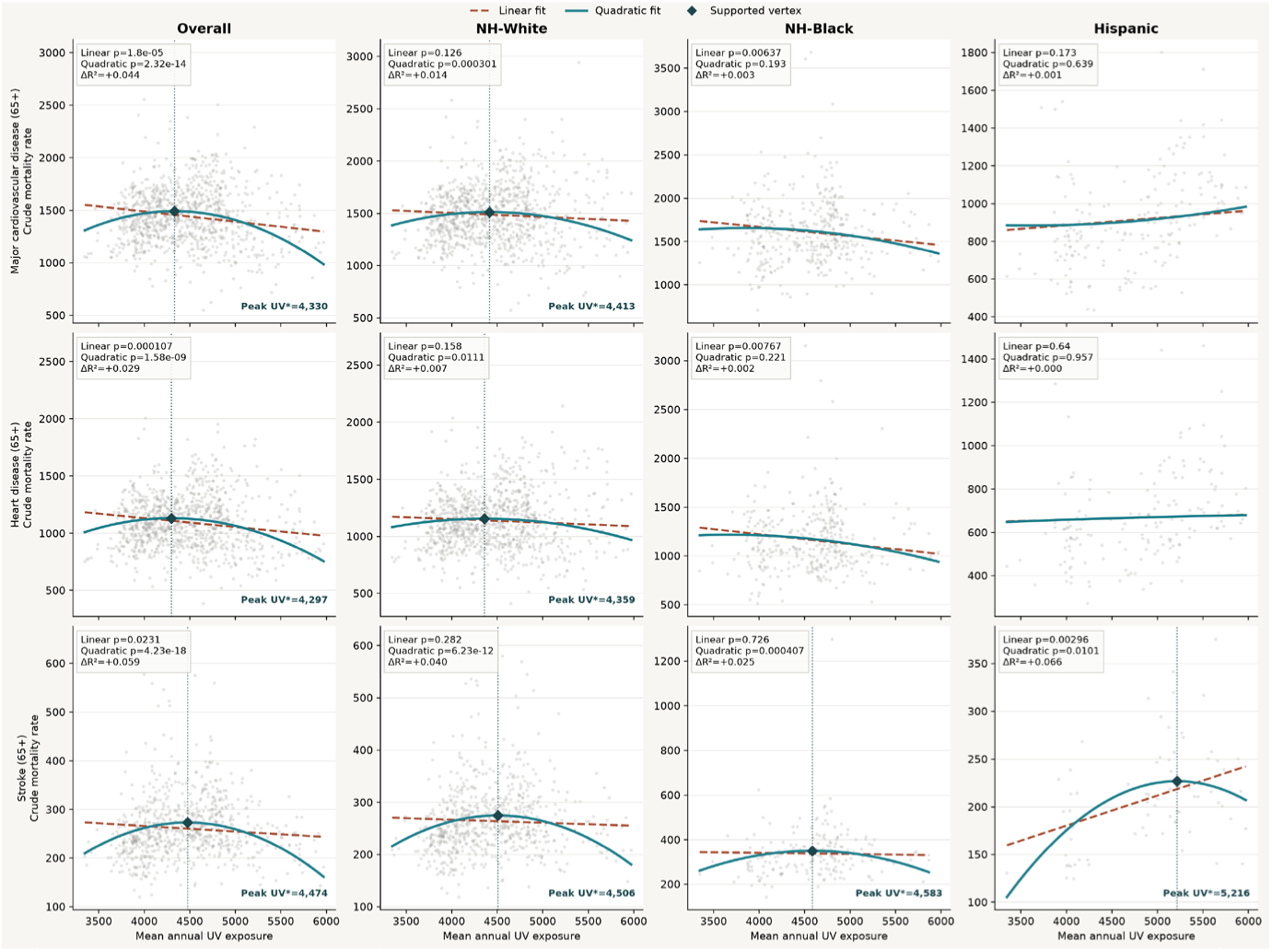
Linear and quadratic fits for ambient ultraviolet exposure and cardiovascular mortality. SVI-adjusted associations are shown for major cardiovascular disease, heart disease, and stroke among US adults aged 65 years or older, overall and by population group. Points represent HSA-level crude mortality rates; dashed lines show linear fits and solid lines show quadratic fits. Curvature primarily reflected a steeper inverse slope in the highest UV quartile rather than attenuation at higher exposure or evidence of an optimal threshold. Diamonds and vertical dotted lines indicate fitted interior turning points, which should not be interpreted as causal thresholds or recommended individual UV exposures. Models used OLS with HC1 heteroskedasticity-robust standard errors. HSA, Health Service Area; OLS, ordinary least squares; SVI, Social Vulnerability Index; UV, ultraviolet.

#### C3. Crude versus age-adjusted sensitivity

HSA-level analyses in the main manuscript used crude mortality rates, because age-adjusted rates are not published at county level for the CDC WONDER Single-Race mortality dataset. Populations differ in age structure, so a descriptive national comparison was used to indicate the likely direction and approximate size of any difference between age-adjusted and crude summaries. Supplementary Table S4 compares the published 2019 national age-adjusted all-cancer mortality rate with the corresponding national crude rate for 2018-2019 used in this paper, overall and by population group. The overall difference is negligible (-0.1%), whereas group-specific differences varied in magnitude and direction: the crude rate is higher than the age-adjusted rate for NH-White adults (-4.6%) and lower for Hispanic (+16.9%) and NH-Black (+8.8%) adults, the latter two differences being substantially larger and consistent with the younger age structure of those populations. These are national descriptive comparisons only and do not provide HSA-level age-adjusted estimates.

**Supplementary Table S4.** National all-cancer mortality rates per 100,000 population: 2019 national age-adjusted rates compared with the 2018-2019 national crude rates used in this paper. Descriptive national comparison only; these are not HSA-level age-adjusted estimates.

| Population group | HSAs | 2019 national age-adjusted rate per 100,000 | Paper 2018-2019 national crude rate per 100,000 | Difference |
| --- | --- | --- | --- | --- |
| All races | 920 | 490.1 | 490.7 | -0.1% |
| NH-White | 914 | 509.1 | 533.6 | -4.6% |
| Hispanic | 198 | 353.1 | 302.0 | +16.9% |
| NH-Black | 375 | 577.6 | 530.8 | +8.8% |

NH, non-Hispanic. Difference is expressed relative to the crude rate, as reported in the approved source output; positive values indicate that the age-adjusted rate exceeds the crude rate and negative values that it is lower. Values are reproduced without recalculation.

##### Limitations of the age-structure analysis

We explored whether HSA mortality rates could be age standardised. CDC WONDER does not provide county-level age-adjusted rates for the Single-Race mortality dataset, so these rates would have to be reconstructed from county counts and populations for each age group.

Suppression made this approach unreliable. Even for lung cancer, the highest-volume individual cancer examined, 57.2% of county-age cells for ages 50-84 and 61.4% of county observations for ages 85 or older were suppressed. Only 7.4% of counties had complete data across all eight age groups. Each suppressed cell contains between one and nine deaths. Counting these cells as zero would underestimate mortality, while excluding or estimating them could bias the results. The problem would be greater for lower-volume cancers and race-specific analyses. We therefore retained crude HSA mortality rates and recognise that differences in population age structure remain a limitation.

#### C4. Melanoma positive-control analysis

Melanoma was already included in the SEER cancer endpoint set and provided a positive control because UV exposure is an established risk factor for melanoma incidence. It also provided a control across race strata because melanoma was sufficiently common for analysis primarily among the NH-White population. A harmful association should therefore be most detectable among NH-White populations, while estimates for NH-Black and Hispanic populations would be limited by low event counts.

Melanoma was the only outcome showing a consistently harmful UV association. This supports the ability of the HSA-level models to detect harmful as well as protective signals.

#### Supplementary Appendix D. Hispanic Cohort and Geographic Sensitivity Analyses

##### Supplementary methods

Since the Hispanic population had the lowest crude mortality rates from both cardiovascular disease and cancer and a unique signature, we conducted a post-hoc analysis restricted to HSAs eligible for the Hispanic analysis. A total of 198 HSAs met the minimum eligibility requirements for the Hispanic cohort. Outcome-specific analyses were conducted within this Hispanic-eligible geographic set, with the number of evaluable HSAs varying by outcome because of event-count, population, and data-availability thresholds; no individual outcome included more than 198 HSAs.

Within this common geographic framework, SVI-adjusted associations between ambient UV and cancer mortality were estimated separately for Hispanic, NH-White, and NH-Black populations using OLS with HC1 heteroskedasticity-robust standard errors. Estimates represent the change in annual crude mortality per 100,000 associated with our previously discussed B UVIQR increase in ambient UV.

The analysis included all-cancers and the individual cancers that were FDR-adjusted for significant in the national all-races analysis. Kidney cancer was borderline significant and was included to examine the Hispanic-specific positive association, and melanoma was retained as the national NH-White positive control. Outcomes becoming significant only within the post-hoc subset were not added. Estimates were not reported when fewer than 10 eligible HSAs remained.

###### D1. National mortality context

The Hispanic population had the lowest crude mortality rates from both cardiovascular disease and cancer. Using 2019 US Census Bureau single-year-of-age population estimates, the population-weighted mean age among residents aged ≥50 years was 65.1 years nationally, 65.8 years among NH-White residents, 63.6 years among NH-Black residents, and 62.6 years among Hispanic residents. Among residents aged ≥65 years, corresponding mean ages were 74.4, 74.6, 73.7, and 74.0 years, respectively. Their younger age may have contributed to this advantage: national age adjustment increased all-cancer mortality from 302.0 to 353.1 deaths per 100,000, while the 65+ difference of 0.4 years younger than the national average is likely not of material impact.^39^ Reliable age-adjusted rates could not be constructed at the HSA level because of extensive suppression of county-age cells, as detailed in Supplementary Appendix C3.

**Supplementary Table S5.** National population averages 2018-2019 by age-group and race-strata in our assessed data sets.

| Population group | Mean age ≥50 (years) | Mean age ≥65 (years) | Population ≥50 / ≥65 |
| --- | --- | --- | --- |
| All races | 65.1 | 74.4 | 117.0 M / 54.1 M |
| NH-White | 65.8 | 74.6 | 83.7 M / 41.2 M |
| NH-Black | 63.6 | 73.7 | 12.5 M / 5.0 M |
| Hispanic | 62.6 | 74.0 | 13.0 M / 4.6 M |

Most Hispanic cancer mortality rates remained below the corresponding all-races rates, with liver and stomach cancer notable exceptions. The Hispanic population also resides, on average, in the highest ambient UV regions of the United States, with mean annual UV exposure approximately 0.4 IQR above the national average(Table 1). This geographic concentration complicated interpretation of the Hispanic UV–mortality findings and motivated the common-geography sensitivity analysis below.

**Supplementary Table S6.**
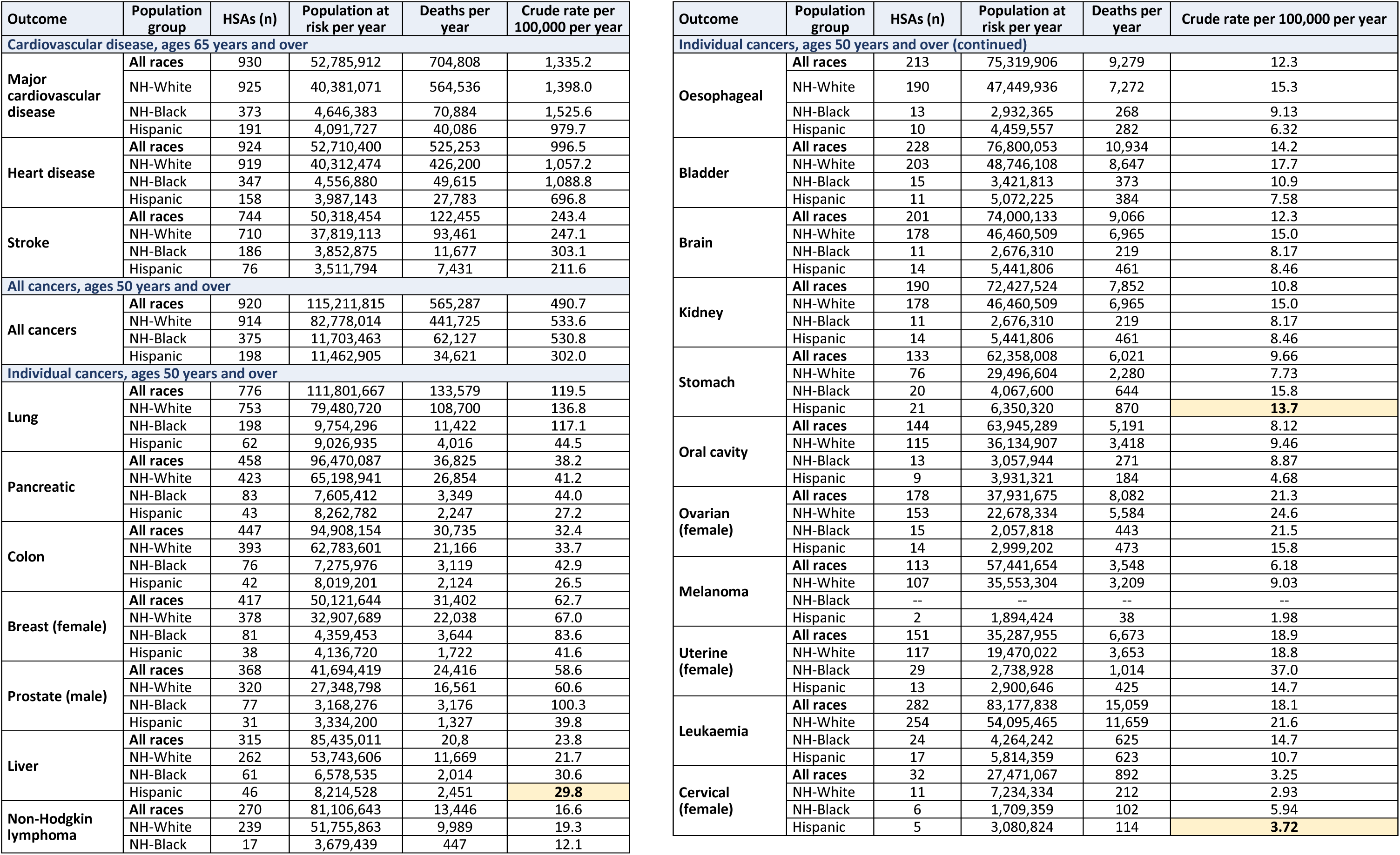
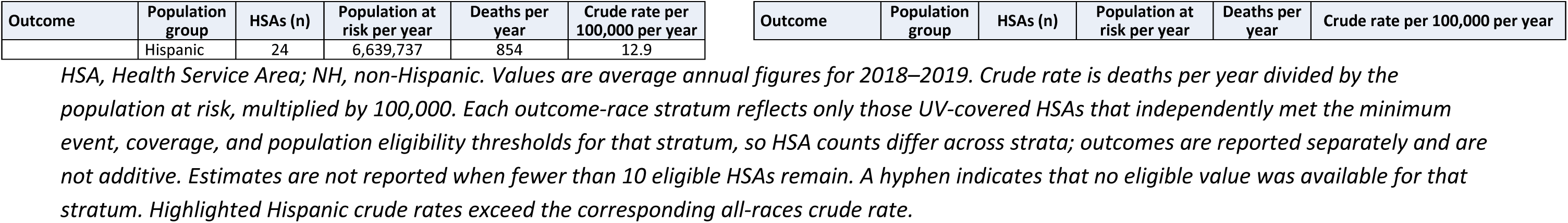
National annual mortality burden and crude mortality rates by race and ethnicity across eligible US Health Service Areas, 2018–2019. Descriptive national context only; values are reproduced from the approved source output without recalculation.

###### D2. Hispanic UV–mortality associations

In analyses of ambient UV and mortality, the Hispanic cohort showed a statistically significant harmful association for all-cancer mortality after Benjamini–Hochberg correction. The sole site-specific positive association was kidney cancer, with an estimated increase of 1.76 β UV/IQR alongside the only NH-Black individually positive association of 3.57 β UV/IQR. Kidney cancer was the only outlier of this direction and magnitude. Melanoma could not be reliably estimated in the Hispanic analysis because fewer than 10 eligible HSAs remained.

Because the Hispanic population is concentrated in higher-UV regions, we considered whether this geographic distribution itself could account for the divergent cancer mortality pattern.

###### D3. Common-geography sensitivity analysis

To test this possibility, we repeated the SVI-adjusted UV-cancer mortality analysis in NH-White and NH-Black populations restricted to the same geographic framework defined by the 198 Hispanic-eligible HSAs. Outcome-specific sample sizes remained subject to the same eligibility requirements and were therefore frequently lower than 198.

No uniformly harmful cancer mortality pattern emerged.

Within this Hispanic-eligible geography, the NH-White population retained a predominantly protective UV-associated mortality signature similar to that observed in the primary national analysis. The NH-Black population, rather than reproducing the Hispanic harmful pattern, shifted from a largely null national profile toward a more protective aggregate all-cancer mortality association.

These findings suggest that broad geographic UV distribution alone does not account for the racial and ethnic heterogeneity observed in the primary analyses. For example, SVI does not fully capture occupational and environmental cancer risks that geographically correspond to high-UV regions. Restricting the comparison populations to the same high-UV geographic framework did not reproduce the Hispanic cancer mortality signature.

**Supplementary Figure S5.**
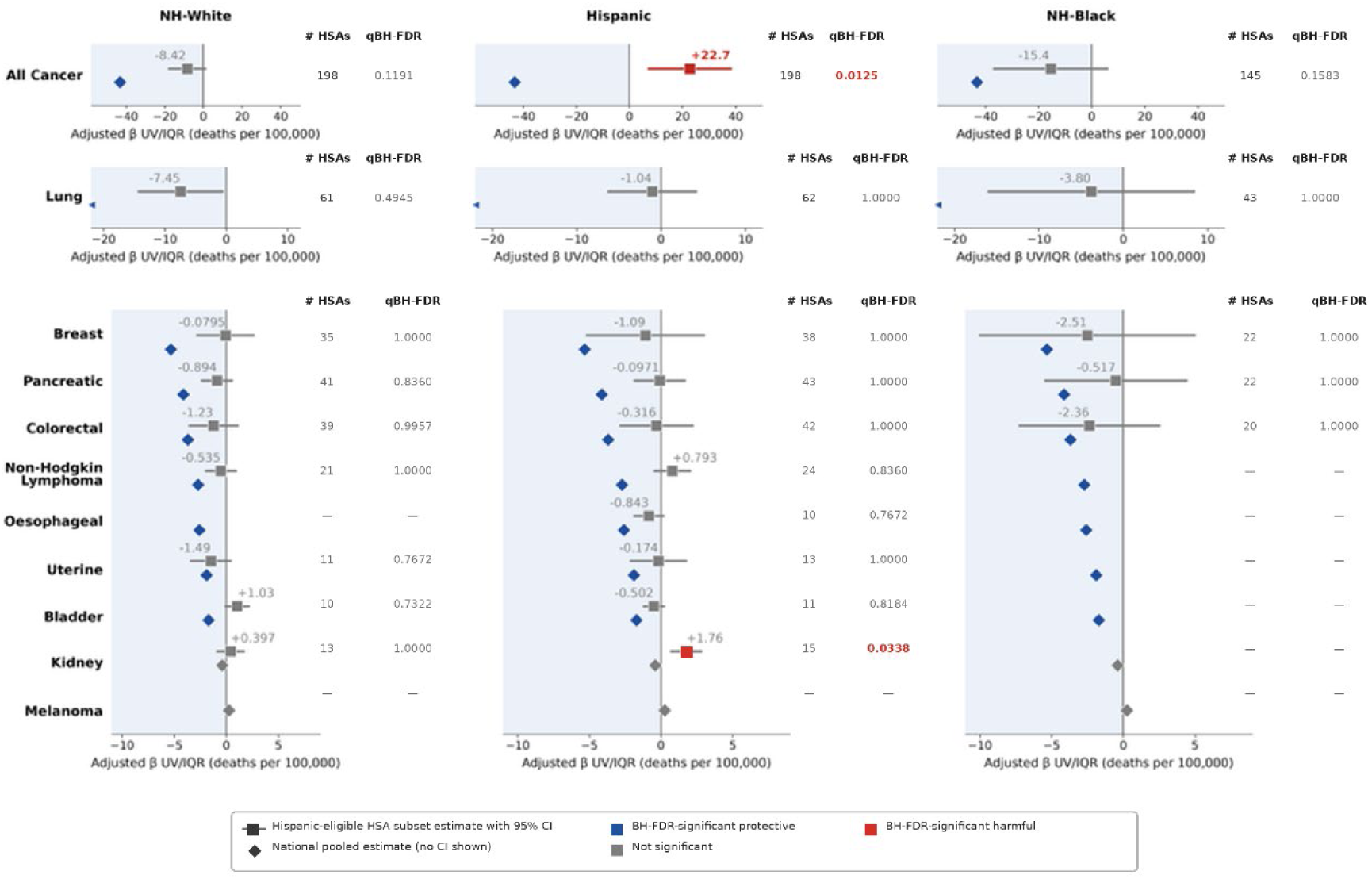
SVI-adjusted associations between ambient ultraviolet exposure and cancer mortality within Hispanic-eligible US Health Service Areas. Estimates for NH-White, Hispanic, and NH-Black populations were calculated within a common set of HSAs eligible for the Hispanic analysis. Squares and horizontal lines represent estimates and 95% confidence intervals; diamonds represent the corresponding national pooled estimates. Blue indicates an FDR-adjusted protective association, grey a non-significant association, and red an FDR-adjusted significant harmful association. Estimates were not reported when fewer than 10 eligible HSAs remained. Notes: Estimates are changes in annual crude mortality per 100,000 associated with an IQR increase in ambient UV and are adjusted for SVI. Outcomes include all cancers, nationally significant individual cancers, kidney cancer as a Hispanic-specific finding, and melanoma as the national NH-White positive control. HSA, Health Service Area; IQR, interquartile range; NH, non-Hispanic; SVI, Social Vulnerability Index; UV, ultraviolet; qBH-FDR, q value of Benjamini–Hochberg false-discovery-rate

###### D4. Potential occupational, environmental, and population modifiers

The Hispanic-specific findings must also be considered in the context of cancer risk factors that are not fully captured by SVI. Hispanic workers comprise more than 75% of the US agricultural workforce,^40^ and agricultural and pesticide-exposed worker cohorts have reported increased risks of stomach, liver, and kidney cancers.^41^ Higher hepatitis prevalence and liver cancer mortality have also been reported among US Hispanic populations independently of occupation.^42,43^

These observations provide context for some of the site-specific differences in the present analysis. In particular, crude Hispanic mortality rates were below the national average for most cancers but not for liver and stomach cancers, while kidney cancer produced the strongest positive UV-associated signal. They do not, however, establish that occupation, pesticide exposure, hepatitis, or any other individual factor explains the Hispanic UV-associated mortality pattern.

Overall, the matched-geography analyses indicate that high ambient UV exposure alone is insufficient to explain the divergent Hispanic cancer mortality signature. Additional occupational, environmental, structural, or biologic modifiers may contribute to the observed heterogeneity and warrant further study.

### Supplementary Appendix E. UK Biobank analysis

#### Supplementary methods: UK Biobank analysis

##### Study population and outcome ascertainment

The UK Biobank analysis used data from participants recruited at baseline assessment centres and followed prospectively through linkage to national cancer registries in England, Wales, and Scotland and to national death records held by NHS England, NHS Wales, and the NHS Central Register for Scotland, which provided cause and date of death. The current analytic cohort comprised 412,665 participants (Supplementary Figure S2).^16^

Two external datasets were available for comparison: one published large-scale US analysis of UV and cancer mortality, and one independent UK Biobank analysis. Both are NH-White cohorts, by definition in the US analysis and, in the UK Biobank, predominantly White Europeans by ancestry. Given our heterogeneous results we chose to compare them to our NH-White cohort. Each cohort measured mortality signals differently.

Individual UV cancer mortality associations from the UK Biobank provided a corroboration dataset.^15^ Cancer-specific mortality from the date of recruitment of 412,665 individuals was assessed with sufficient data, applying a fully adjusted Cox proportional hazards model, including age, sex, smoking, physical activity, employment, deprivation, education, BMI, and solarium use (Supplementary Table S7). The reported hazard ratios represent an estimated change in mortality risk associated with a 2,000 kJ/m² higher residential SWR (SWR, 2008).^15^

We conducted complementary analyses in 412,665 UK Biobank participants, an analytic sample defined for the present cause-specific outcome set and therefore distinct from the 395,086 complete-case sample of the previously published mortality analysis, extending previously reported associations between residential shortwave radiation and mortality to heart disease, stroke, and site-specific cancer outcomes.^15^ Cause-specific mortality was ascertained through linked national registry and NHS death records. Annual mean residential shortwave radiation was assigned from 2008 satellite estimates and modelled per 2000 kJ/m² increase.

Cox proportional hazards models estimated hazard ratios and 95% confidence intervals from recruitment. Age-adjusted and fully adjusted models included age, sex where applicable, smoking, physical activity, employment, education, area-level deprivation, body mass index, and sunbed use. Cancer analyses were restricted to participants of White British or other White ancestry, with sex omitted from sex-specific cancer models. Benjamini-Hochberg false discovery rate adjustment was applied within the family of individual cancer outcomes.25 Full cohort, exposure, outcome, and analytical methods are provided in the Supplementary Methods; estimates are reported in Supplementary Table S7 and Supplementary Figure S6.

Outcomes were cause-specific mortality from heart disease and from stroke; two pooled cancer outcomes (all-cancer mortality and non-skin cancer mortality); and 20 individual cancer outcomes (lung, bowel, breast, pancreatic, prostate, oesophageal, brain, liver, NHL, ovarian, leukaemia, kidney, stomach, bladder, mesothelioma, melanoma, uterine, oral, thyroid, and cervix). Cause-specific deaths were identified from the underlying (primary) cause of death recorded in the death registration and coded according to the International Classification of Diseases, 10th Revision (ICD-10).

The number of deaths varied by outcome and is reported for each model in Supplementary Table S7. The cohort denominator and outcome-specific event counts should therefore be interpreted separately.

Survival time was calculated in person-years from the date of baseline assessment (recruitment) to the date of the relevant cause-specific death, loss to follow-up, or the end of the study period, whichever occurred first.

##### Residential shortwave radiation exposure

The exposure was annual mean residential downward shortwave radiation (250-2500 nm), expressed in kJ/m². Monthly downward shortwave radiation estimates covering the UK on a 0.05° x 0.05° grid were obtained from the Japan Aerospace Exploration Agency (JAXA) and were derived from the Moderate Resolution Imaging Spectroradiometer (MODIS) instrument aboard NASA’s Aqua and Terra satellites, accounting for cloud and aerosol thickness. These estimates span the ultraviolet, visible, near-infrared, and infrared portions of the spectrum.

An annual average for 2008 was calculated from the monthly grid-point values and used as the exposure metric, 2008 being close to the midpoint of the UK Biobank recruitment period. Participants’ residential locations, supplied as Ordnance Survey National Grid coordinates at 1 km x 1 km resolution, were converted to longitude and latitude and spatially joined to the nearest shortwave radiation estimate. The exposure was rescaled so that each hazard ratio corresponds to a 2000-unit increase in annual mean residential shortwave radiation.

##### Covariates

Covariates were selected a priori as factors plausibly associated with both residential shortwave radiation and mortality. They comprised age at recruitment; sex; employment status (employed or unemployed); education (completion of a college or university-level degree); area-level deprivation, measured by a UK-adjusted Index of Multiple Deprivation treated as a continuous variable and rescaled across England, Wales, and Scotland; smoking status (never, or former or current); vigorous physical activity (none, or one or more days per week lasting more than 10 minutes); body mass index category (below 25, 25-30, or 30 kg/m² or above); and recent recreational sunbed use.

Sex was omitted from models for cancers occurring in a single sex (prostate, breast, uterine, ovary, and cervix). Sex was included as a covariate in the corrected fully adjusted cardiovascular models reported in Supplementary Table S7.

#### Statistical analysis

All analyses were performed in Stata 18.0 (StataCorp). Descriptive statistics were summarised as means and standard deviations for continuous variables and as proportions for categorical variables. Cox proportional hazards models were fitted separately for each outcome to estimate hazard ratios and 95% confidence intervals for the association between annual mean residential shortwave radiation and cause-specific mortality from recruitment onwards. For each outcome, two specifications were fitted: an age-adjusted model, including age at recruitment, and a fully adjusted model, including all covariates listed above.

Cancer analyses were restricted to participants of white British, related white British, or white non-British ancestry. Cardiovascular analyses (heart disease and stroke mortality) are reported alongside the cancer models in Supplementary Table S7.

Benjamini-Hochberg false discovery rate adjustment was applied within the family of 20 individual cancer outcomes. Pooled cancer and cardiovascular outcomes were excluded from this correction. FDR-adjusted values for the individual cancer outcomes are reported in Supplementary Table S7.

Estimates for all outcomes and both model specifications are presented in Supplementary Table S7 and are displayed graphically, for both the age-adjusted and fully adjusted models, in Supplementary Figure S6.

##### Ethics

UK Biobank data were accessed and analysed under UK Biobank Application 82526. UK Biobank holds ethical approval from the Northwest Multi-centre Research Ethics Committee (REC reference 11/NW/0382), and all participants provided written informed consent at enrolment. The present work was a secondary analysis of de-identified data obtained from the UK Biobank resource.

**Supplementary Table S7.**
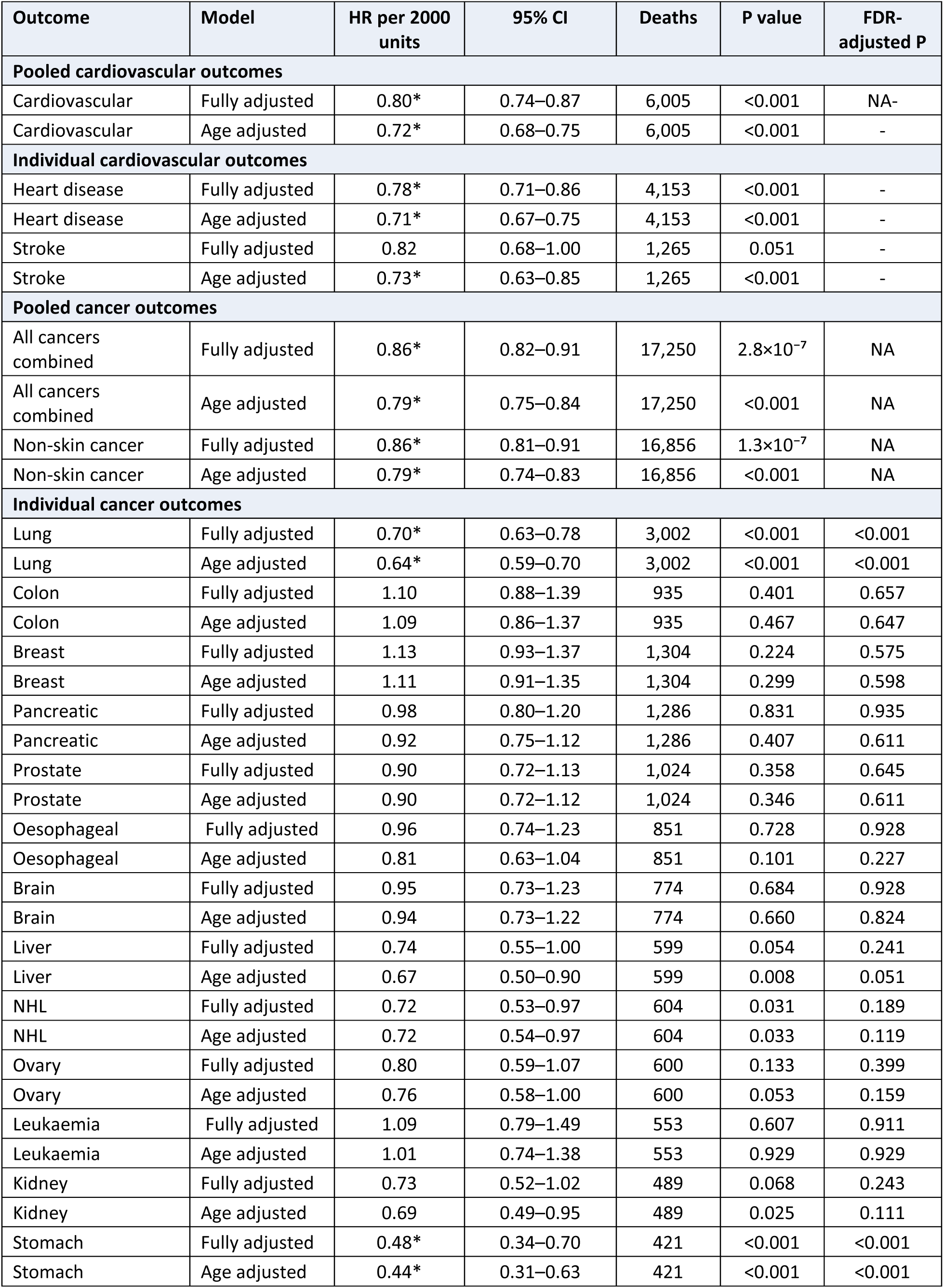
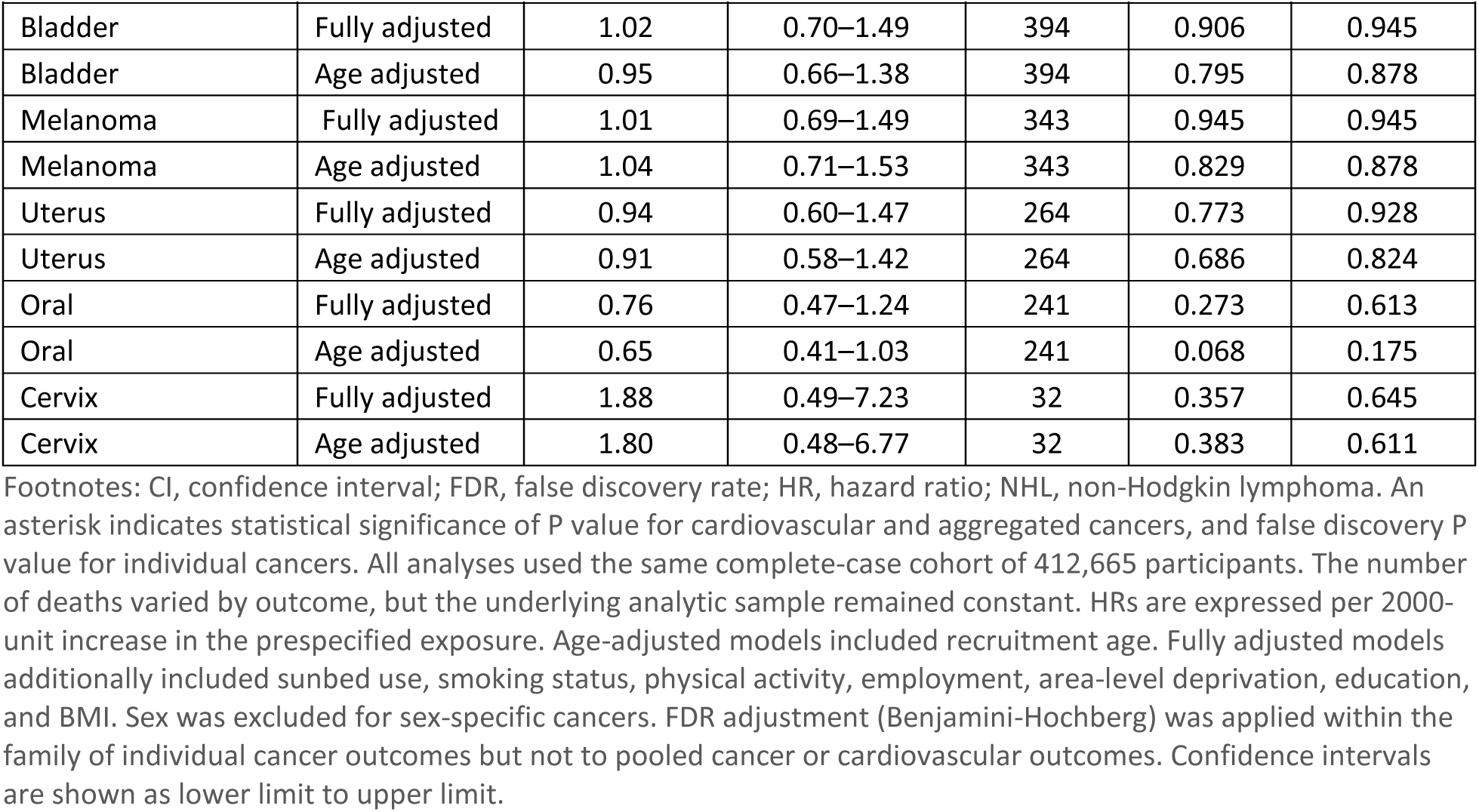
Associations between exposure and cause-specific mortality in 412,665 UK Biobank participants.

| Outcome | Model | HR per 2000 units | 95% CI | Deaths | P value | FDR-adjusted P |
| --- | --- | --- | --- | --- | --- | --- |
| **Pooled cardiovascular outcomes** |  |  |  |  |  |  |
| Cardiovascular | Fully adjusted | 0.80* | 0.74–0.87 | 6,005 | <0.001 | NA- |
| Cardiovascular | Age adjusted | 0.72* | 0.68–0.75 | 6,005 | <0.001 | - |
| **Individual cardiovascular outcomes** |  |  |  |  |  |  |
| Heart disease | Fully adjusted | 0.78* | 0.71–0.86 | 4,153 | <0.001 | - |
| Heart disease | Age adjusted | 0.71* | 0.67–0.75 | 4,153 | <0.001 | - |
| Stroke | Fully adjusted | 0.82 | 0.68–1.00 | 1,265 | 0.051 | - |
| Stroke | Age adjusted | 0.73* | 0.63–0.85 | 1,265 | <0.001 | - |
| **Pooled cancer outcomes** |  |  |  |  |  |  |
| All cancers combined | Fully adjusted | 0.86* | 0.82–0.91 | 17,250 | 2.8×10<sup>-7</sup> | NA |
| All cancers combined | Age adjusted | 0.79* | 0.75–0.84 | 17,250 | <0.001 | NA |
| Non-skin cancer | Fully adjusted | 0.86* | 0.81–0.91 | 16,856 | 1.3×10<sup>-7</sup> | NA |
| Non-skin cancer | Age adjusted | 0.79* | 0.74–0.83 | 16,856 | <0.001 | NA |
| **Individual cancer outcomes** |  |  |  |  |  |  |
| Lung | Fully adjusted | 0.70* | 0.63–0.78 | 3,002 | <0.001 | <0.001 |
| Lung | Age adjusted | 0.64* | 0.59–0.70 | 3,002 | <0.001 | <0.001 |
| Colon | Fully adjusted | 1.10 | 0.88–1.39 | 935 | 0.401 | 0.657 |
| Colon | Age adjusted | 1.09 | 0.86–1.37 | 935 | 0.467 | 0.647 |
| Breast | Fully adjusted | 1.13 | 0.93–1.37 | 1,304 | 0.224 | 0.575 |
| Breast | Age adjusted | 1.11 | 0.91–1.35 | 1,304 | 0.299 | 0.598 |
| Pancreatic | Fully adjusted | 0.98 | 0.80–1.20 | 1,286 | 0.831 | 0.935 |
| Pancreatic | Age adjusted | 0.92 | 0.75–1.12 | 1,286 | 0.407 | 0.611 |
| Prostate | Fully adjusted | 0.90 | 0.72–1.13 | 1,024 | 0.358 | 0.645 |
| Prostate | Age adjusted | 0.90 | 0.72–1.12 | 1,024 | 0.346 | 0.611 |
| Oesophageal | Fully adjusted | 0.96 | 0.74–1.23 | 851 | 0.728 | 0.928 |
| Oesophageal | Age adjusted | 0.81 | 0.63–1.04 | 851 | 0.101 | 0.227 |
| Brain | Fully adjusted | 0.95 | 0.73–1.23 | 774 | 0.684 | 0.928 |
| Brain | Age adjusted | 0.94 | 0.73–1.22 | 774 | 0.660 | 0.824 |
| Liver | Fully adjusted | 0.74 | 0.55–1.00 | 599 | 0.054 | 0.241 |
| Liver | Age adjusted | 0.67 | 0.50–0.90 | 599 | 0.008 | 0.051 |
| NHL | Fully adjusted | 0.72 | 0.53–0.97 | 604 | 0.031 | 0.189 |
| NHL | Age adjusted | 0.72 | 0.54–0.97 | 604 | 0.033 | 0.119 |
| Ovary | Fully adjusted | 0.80 | 0.59–1.07 | 600 | 0.133 | 0.399 |
| Ovary | Age adjusted | 0.76 | 0.58–1.00 | 600 | 0.053 | 0.159 |
| Leukaemia | Fully adjusted | 1.09 | 0.79–1.49 | 553 | 0.607 | 0.911 |
| Leukaemia | Age adjusted | 1.01 | 0.74–1.38 | 553 | 0.929 | 0.929 |
| Kidney | Fully adjusted | 0.73 | 0.52–1.02 | 489 | 0.068 | 0.243 |
| Kidney | Age adjusted | 0.69 | 0.49–0.95 | 489 | 0.025 | 0.111 |
| Stomach | Fully adjusted | 0.48* | 0.34–0.70 | 421 | <0.001 | <0.001 |
| Stomach | Age adjusted | 0.44* | 0.31–0.63 | 421 | <0.001 | <0.001 |
| Bladder | Fully adjusted | 1.02 | 0.70–1.49 | 394 | 0.906 | 0.945 |
| Bladder | Age adjusted | 0.95 | 0.66–1.38 | 394 | 0.795 | 0.878 |
| Melanoma | Fully adjusted | 1.01 | 0.69–1.49 | 343 | 0.945 | 0.945 |
| Melanoma | Age adjusted | 1.04 | 0.71–1.53 | 343 | 0.829 | 0.878 |
| Uterus | Fully adjusted | 0.94 | 0.60–1.47 | 264 | 0.773 | 0.928 |
| Uterus | Age adjusted | 0.91 | 0.58–1.42 | 264 | 0.686 | 0.824 |
| Oral | Fully adjusted | 0.76 | 0.47–1.24 | 241 | 0.273 | 0.613 |
| Oral | Age adjusted | 0.65 | 0.41–1.03 | 241 | 0.068 | 0.175 |
| Cervix | Fully adjusted | 1.88 | 0.49–7.23 | 32 | 0.357 | 0.645 |
| Cervix | Age adjusted | 1.80 | 0.48–6.77 | 32 | 0.383 | 0.611 |

Footnotes: CI, confidence interval; FDR, false discovery rate; HR, hazard ratio; NHL, non-Hodgkin lymphoma. An asterisk indicates statistical significance of P value for cardiovascular and aggregated cancers, and false discovery P value for individual cancers. All analyses used the same complete-case cohort of 412,665 participants. The number of deaths varied by outcome, but the underlying analytic sample remained constant. HRs are expressed per 2000-unit increase in the prespecified exposure. Age-adjusted models included recruitment age. Fully adjusted models additionally included sunbed use, smoking status, physical activity, employment, area-level deprivation, education, and BMI. Sex was excluded for sex-specific cancers. FDR adjustment (Benjamini-Hochberg) was applied within the family of individual cancer outcomes but not to pooled cancer or cardiovascular outcomes. Confidence intervals are shown as lower limit to upper limit.

**Supplementary Figure S6.**
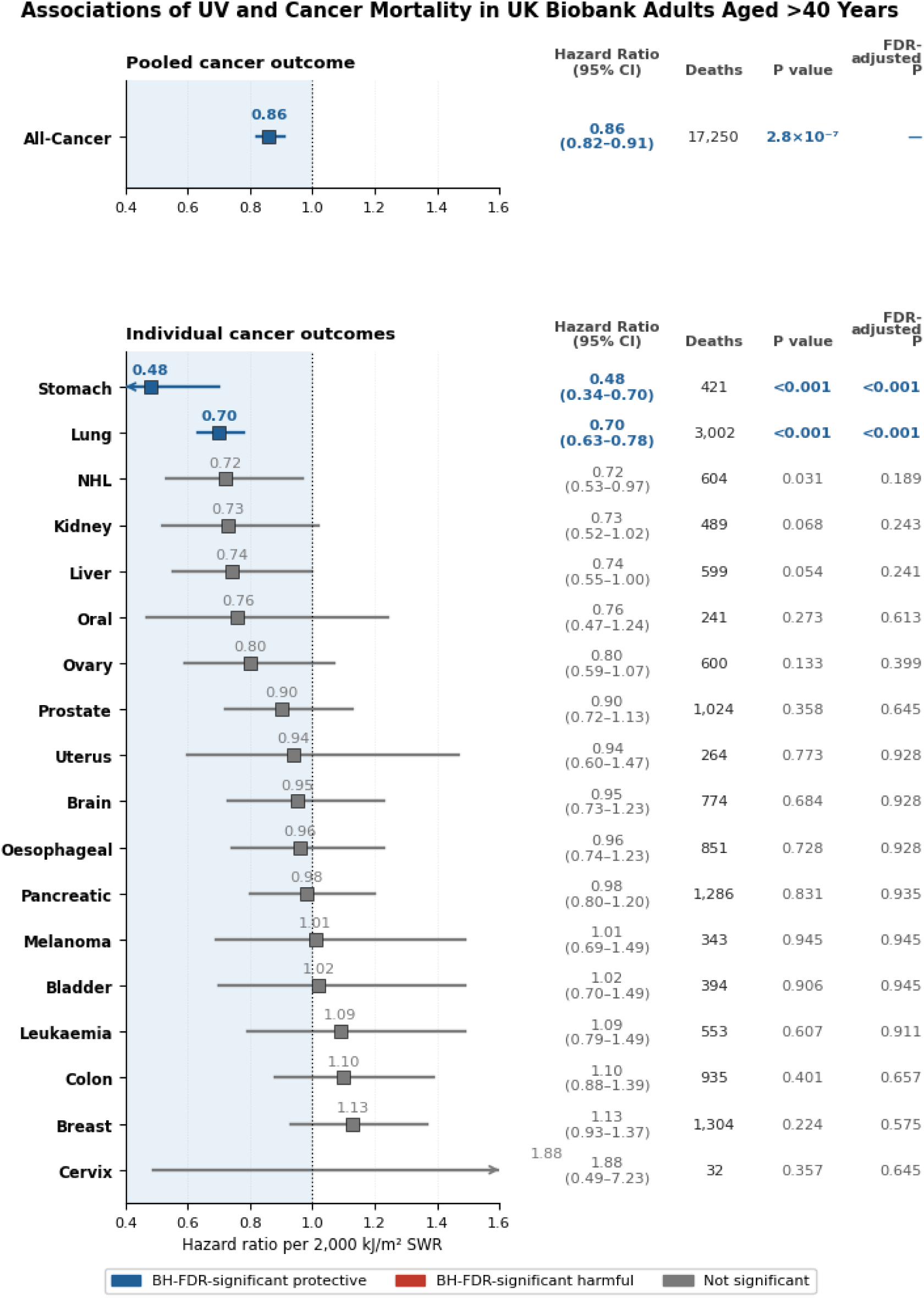
UK Biobank associations between exposure and cause-specific mortality by fully adjusted model. Points represent hazard ratios (HRs) per 2000 kJ/m^2^ SWR increase in exposure; horizontal lines represent 95% confidence intervals. Filled circles indicate fully adjusted estimates and open squares indicate age-adjusted estimates. The vertical reference line denotes HR = 1.0. Values below 1 indicate lower mortality. The plotting range is restricted to 0.4-1.6 for readability; arrowheads indicate confidence intervals extending beyond the displayed range. The same numerical estimates, along with age-adjusted models, are reported in Supplementary Table S7.

### Supplementary Appendix F. Proposed exposure-pattern model of UV-associated mortality

Ambient UV exposure was associated in our ecological analyses with lower cardiovascular mortality and mortality from selected cancers. We consider three biologically grounded pathways that may contribute to this pattern: (1) vascular responses supported by controlled human exposure studies; (2) photoimmune responses supported by experimental research and clinical phototherapy literature; and (3) a cancer-surveillance hypothesis in which recurrent nonerythemal UV exposure may influence host– tumour interactions. Studies of narrowband UVB (NB-UVB) provide a clinically standardised model^44^ for characterising selected UVB-mediated vascular, immune, and cellular responses. Although NB-UVB treatment is not identical to ambient solar exposure, it offers a controlled human framework for examining mechanisms potentially relevant to habitual nonburning UV exposure.

The observed associations of ambient UV exposure with cardiovascular mortality and mortality from multiple cancer sites require a biologically coherent explanation. How can UV exposure be associated both with increased UV-signature mutation burden,^45^ in exposed skin and with lower inflammatory activity and lower mortality from selected cancers? The central question is therefore not whether UV exposure is uniformly beneficial or harmful, but whether its biological consequences differ according to exposure pattern, tissue context, and the balance between photodamage and recovery.

The cardiovascular pathway involves cutaneous nitric-oxide mobilisation and downstream vascular effects, supported by controlled exposure studies and clinical observations. The photoimmune pathway involves systemic modulation of inflammatory and immune signalling and is well characterised in experimental research and in the context of NB-UVB therapy. A third pathway, cancer surveillance, is considered as a hypothesis: recurrent nonerythemal UV exposure may influence cellular stress responses and host–tumour interactions, including apoptosis, antigen processing, and immune recognition of abnormal cells. Evidence from clinical research and long-term follow-up of NB-UVB– treated populations is reviewed for mechanistic insight. We then examine whether convergence of these pathways may help explain why the inverse association is concentrated in selected cancers rather than observed uniformly across all cancer sites. Finally, we present an exposure-pattern model distinguishing regular nonburning exposure from intermittent burning exposure.

We consider three pathways. The cardiovascular pathway, supported by experimental and clinical evidence, involves cutaneous nitric-oxide mobilisation and downstream vascular effects. The photoimmune pathway, also well established in the context of NB-UVB therapy, involves systemic modulation of inflammatory and immune signalling. A third pathway, cancer surveillance, is considered as a hypothesis: recurrent nonerythemal UV exposure may influence cellular stress responses and host– tumour interactions, including apoptosis, antigen processing, and adaptive immune priming or T-cell memory.^46,47^ Evidence from clinical research and long-term follow-up of NB-UVB–treated populations is reviewed for mechanistic insight. We then examine whether convergence of these pathways may help explain why the inverse association is concentrated in selected cancers rather than observed uniformly across all cancer sites. Finally, we present an exposure-pattern model distinguishing regular nonburning exposure from intermittent burning exposure.

#### Cardiovascular Pathway

UV exposure activates a documented cardiovascular pathway through the release of nitric oxide and related vascular signalling. These responses improve vascular tone, reduce blood pressure, and reduce vascular inflammation. Evidence from environmental studies, controlled exposure research, and randomised NB-UVB trials supports effects beyond the skin, including lower systolic blood pressure (SBP), lower high-sensitivity C-reactive protein (hs-CRP) and interleukin-6 (IL-6), and increased high-density lipoprotein cholesterol (HDL-C).^48–50^ Given all are recognised predictors of CVD mortality by the American College of Cardiology,^51^ this pathway provides the most direct biological explanation for the strong association observed between ambient UV and lower CVD mortality, particularly the dominant heart disease signal.

These acute physiological findings are complemented by longer-term clinical observations among patients treated with NB-UVB. In an 11-year nationwide propensity-score–matched cohort, Bae et al. compared 3,229 patients with vitiligo who received at least 100 NB-UVB treatments with 9,687 who received fewer than three treatments.^52^ The incidence of combined cardiovascular and cerebrovascular events was 60.0 versus 95.6 per 10,000 person-years, corresponding to a lower hazard among those receiving at least 100 treatments (HR 0.637).^53^ Cardiovascular events were lower (HR 0.682), as were cerebrovascular events (HR 0.601), including ischaemic heart disease and cerebral infarction.^52,54^

The cohort findings are directionally consistent with the vascular mechanism demonstrated in controlled exposure studies and with the inverse cardiovascular associations observed in the present ecological analysis. Because treatment allocation was not random, however, the results should be interpreted as supportive clinical evidence rather than as proof that NB-UVB treatment caused the lower event rates.

The cardiovascular pathway alone is unlikely to account for the inverse associations observed across several inflammation-linked cancer sites. A second pathway therefore concerns UV-mediated modulation of systemic immune and inflammatory responses.

#### Photoimmune Pathway

The photoimmune pathway is well characterised in experimental research and in the clinical use of narrowband ultraviolet B (NB-UVB) phototherapy. Cutaneous UV exposure alters epidermal and immune-cell signalling; dendritic-cell responses can shape downstream adaptive immunity,^55^ with reported effects including increased FoxP3+ regulatory T cells, reduced inflammatory signalling, and shifts towards immune homeostasis.^56,57^ Human studies also document UV-associated changes in circulating immune-cell number, phenotype, and function, supporting systemic effects beyond irradiated skin.^58–60^

Clinically, NB-UVB has been used for decades to treat inflammatory and immune-mediated diseases, including psoriasis, graft-versus-host disease, vitiligo, and early multiple sclerosis.^44,61–63^ At prescribed suberythemal doses, it is best understood as context-dependent immune regulation rather than uniform immunosuppression. In a controlled comparison of psoriasis treatments, NB-UVB increased circulating and cutaneous FoxP3+ regulatory T cells, reduced Th1 cells, and the rise in cutaneous regulatory T cells correlated with clinical improvement.^64^ This comparison supports a photoimmune effect not explained solely by vitamin D.

A foundational demonstration of systemic photoimmunology came from murine tumour-transplant experiments in the 1970s. UV-induced tumours were rejected by nonirradiated syngeneic mice but grew in mice previously exposed to UV, demonstrating that cutaneous UV exposure can modify systemic immune responses relevant to tumour rejection.^65^ The experiment established that UV can affect host immunity beyond irradiated skin, providing an important basis for considering how exposure pattern may influence host–tumour interactions.

UV exposure may also produce longer-lasting effects on innate immune function. BCG-induced trained immunity illustrates how metabolic and epigenetic reprogramming of monocytes can generate durable functional responses beyond the lifespan of individual circulating cells.^66,67^ In murine bone-marrow-chimera experiments, prior UV exposure altered dendritic-cell progenitors and the immune-priming capacity of their differentiated progeny after long-term engraftment, consistent with persistent myeloid reprogramming.^68–74^ The UV phenotype differs from canonical BCG-associated trained immunity, but together these findings show that transient environmental or immunological stimuli can durably reprogramme bone-marrow-derived innate immune cells. This provides a mechanistic basis for systemic UV-associated modulation of inflammatory and immune signalling over time.

#### Cancer Surveillance Pathway

Transformative medical insights often emerge from conditions that expose the limits of an accepted model. Early-stage mycosis fungoides (CTCL) and vitiligo provide such an opportunity. As a controlled surrogate for ambient UV exposure, NB-UVB offers an accidental human experiment-and a unique window through which to examine whether the environmental UV signal at the centre of this paper contributes causally to cancer surveillance and systemic resilience.

We show that cancer mortality reduction is most highly correlated with the NH-White population, and we demonstrate significant correlations across multiple types of cancer. Given darker skin inhibit vitamin D photosynthesis, could it be possible that the NH-Black population does not producing enough vitamin D from sunlight to reduce the relative risk of fatal cancer, even in high exposure areas? Would an even higher amount of UV exposure be necessary for this cohort to see a reduction in relative risk?

Another possible hypothesis is that the increased mutation burden associated with UV exposure elicits protective immune responses to cancer. This was first proposed in a 1937 study in which Peller first recognised overall cancer mortality reduction despite higher incidence of skin cancer in sailors.^1^ Despite not having an understanding of adaptive immune mechanisms, or cancer mutagenesis, Peller proposed that the same biological effect of sun exposure driving skin cancer formation provided protection from internal cancers. Now we understand that UV exposure induces coordinated gene-regulatory responses, including changes in microRNA expression,^75^ DNA repair, apoptosis, antigen presentation, and inflammatory signalling, providing a plausible molecular mechanism through which repeated low-level UV exposure could shape adaptive immune responses. Intermittent, sublethal UV-induced damage could be analogous to a nonspecific, “vaccine-like” stimulus by expanding the repertoire of MHC-presented antigens generated by low-level UV-induced mutations in protein-coding sequences, and activating T-cells corresponding to tumour associated antigens. Studies with NB-UVB therapy for psoriasis hint at this mechanism. For CTCL cancer, UV is therapeutic rather than merely tolerated. It is presently a guideline-directed, first-line skin-directed therapy for early-stage mycosis fungoides, the most common form of cutaneous T-cell lymphoma, where it can induce objective and complete clinical responses.^76^ In patients with psoriasis, a single treatment course increased mutation burden by 1.16-to 10.5-fold in infrequently sun-exposed skin and by up to 2.33-fold in frequently exposed skin.^45^ Yet this measurable genomic stress has not been matched by a proportionate increase in malignancy. In a nationwide cohort of 60,321 patients with vitiligo, NB-UVB was not associated with increased melanoma or nonmelanoma skin cancer, including among 717 patients receiving at least 500 treatments.

Furthermore, in a 16-year nationwide matched cohort of 13,824 patients with vitiligo and 55,296 controls, among patients receiving more than 100 phototherapy sessions, unadjusted cancer incidence was 65% lower than in similarly stratified controls.^53^ It is worth considering that UV exposure may be driving adaptive immune responses that are protective to cancer, and allogeneic T cell transfer in mice would be an attractive method to study this effect.

#### Convergence of the Pathways

The cardiovascular and photoimmune pathways provide a coherent biological framework for the inverse associations observed with cardiovascular mortality and mortality from selected cancer sites. The cancer pattern was site-specific rather than uniform: associations were strongest for lung, breast, pancreatic, colon, non-Hodgkin lymphoma, oesophageal, prostate, uterine, and bladder cancers, whereas other sites showed weak or inconsistent associations.

These cancers differ in tissue of origin and proximal aetiology, yet many are shaped by a common set of cancer hallmark processes: chronic inflammation, immune-cell recruitment and evasion, metabolic dysregulation, vascular remodelling, and reciprocal signalling between malignant, stromal, and immune cells.^46,47^ The proposed UV-responsive pathways intersect with these processes. Nitric-oxide mobilisation provides a vascular mechanism; photoimmune regulation influences inflammatory resolution and systemic immune signalling; and recurrent nonerythemal UV-induced cellular stress may promote apoptosis, antigen processing, adaptive immune priming, and immune recognition of abnormal cells.

The convergence of these pathways offers an explanation for why the association is concentrated in particular cancer sites rather than distributed uniformly across all malignancies. Cancers with a greater dependence on inflammatory, vascular, and immune microenvironmental processes may be especially responsive to a recurring environmental stimulus that influences those systems. Most of the cancers showing a protective UV association here, colon, lung, and breast (wound-healing subtype C1) and pancreatic (inflammatory subtype C3), fall within Thorsson’s inflammation and wound-healing-driven immune subtypes, in which an active but incompletely resolved immune response shapes prognosis, consistent with a role for UV-associated inflammatory resolution. This framework leads directly to the exposure-pattern model in Supplementary Figure S7: regular nonburning UV is proposed to support recurring vascular, immune, and cellular-adaptive responses, whereas intermittent burning exposure produces a distinct pattern dominated by photodamage.

Lung cancer provides the clearest example of this convergence. It was the dominant protective signal in both our US analysis and the UK Biobank mortality analysis. Pandya et al. recently identified a 14-protein plasma signature that predicted lung cancer more than five years before diagnosis and was validated across eight cohorts.^77^ Experimental analyses indicated that tumour-promoting inflammatory factors converged on an alveolar transitional state implicated in lung adenocarcinoma initiation and independent NB-UVB research has stabilized broad proteomic panels.^78^ This provides a specific biological setting in which the photoimmune and cancer-surveillance pathways proposed in our model may overlap. Because the Pandya participants are nested within the same UK Biobank population used by Stevenson et al., applying Stevenson’s active and passive UV-exposure measures to the Pandya cohort could directly test whether greater UV exposure is associated with attenuation of this inflammatory signature, reduced progression to lung cancer, or both.

In contrast, cervical, oral, brain, and kidney cancers showed weak, null, or inconsistent UV associations in our data, with similarly limited consistency in the UK Biobank results. These cancers do not lack the hallmarks of cancer, but their dominant aetiologies and tissue environments may be less directly modified by the pathways proposed here.

#### The Proposed Exposure-Pattern Model

This model does not dispute the harms of intermittent burning UV exposure but reconciling the epidemiological and clinical data necessitates challenging the assumption that all UV exposure is biologically equivalent or uniformly harmful. Our integrated model therefore proposes that regular nonburning UV provides a recurring biological stimulus across vascular, immune, and cancer-surveillance systems. The pathways are distinct but synergistic: cardiovascular signalling improves vascular tone and blood pressure, photoimmune signalling promotes inflammatory resolution and immune homeostasis, and neo-epitope generation strengthens early recognition of abnormal cells. Their convergence may explain the reproducible signal across CVD and a subset of inflammation-linked cancers, rather than a uniform effect across all malignancies. The balance between stress, recovery, and adaptation determines whether UV contributes to resilience or harm (Figure S7).

Regular nonburning UV is proposed to provide a recurring biological stimulus across three interacting pathways: vascular nitric-oxide signalling, photoimmune and inflammatory regulation, and cancer surveillance through DNA repair, apoptosis, antigen presentation, and immune recognition. Repeated cycles of stress, recovery, and adaptation may strengthen these responses and contribute to lower cardiovascular and cancer mortality.

**Supplementary Figure S7.**
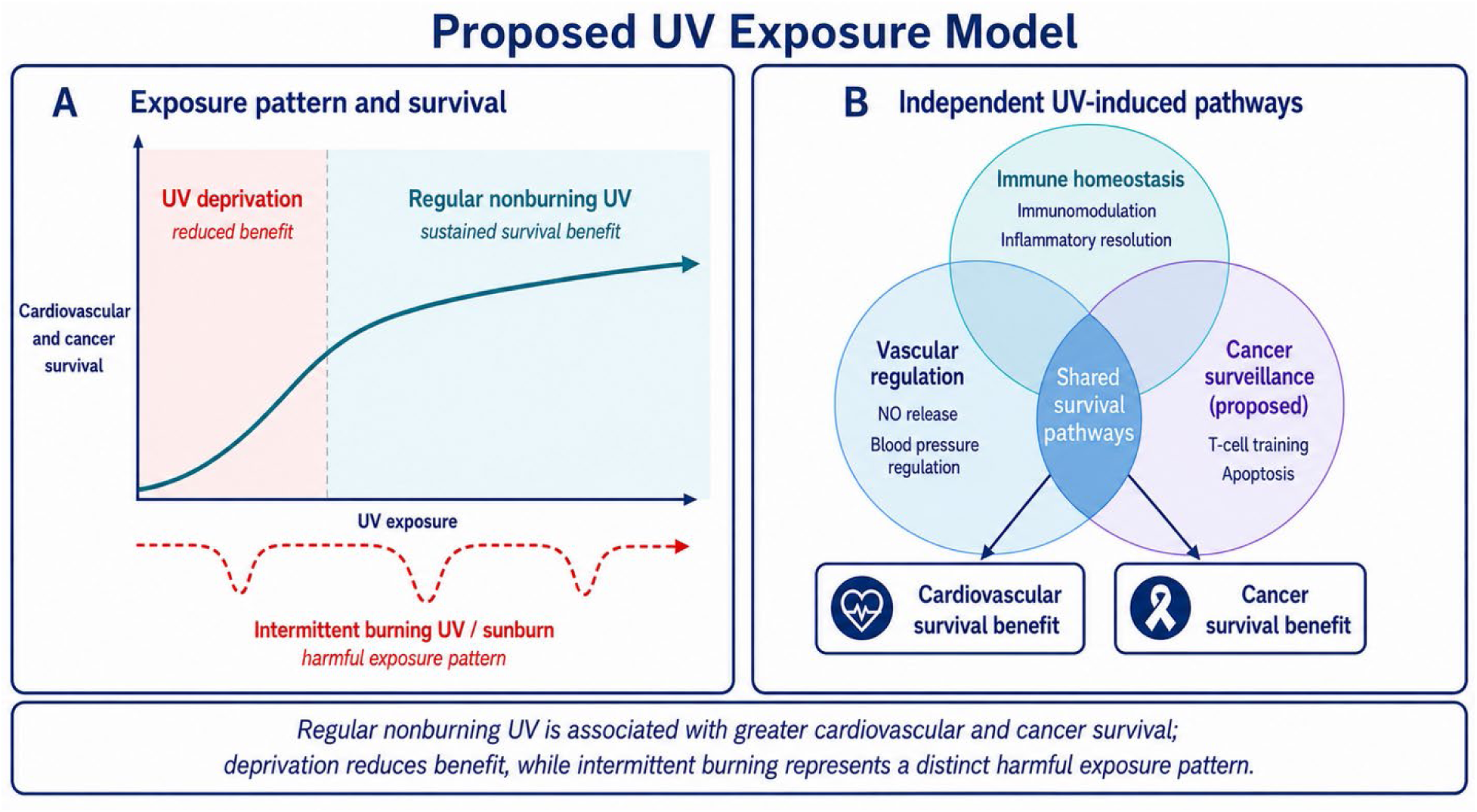
Proposed exposure-pattern model of ultraviolet exposure and mortality across vascular, photoimmune, and cancer-surveillance pathways. Regular nonburning UV provides a recurring stimulus for stress, recovery, and adaptation across three interacting biological systems. Their combined effects offer a unifying explanation for the lower cardiovascular and selected cancer mortality associated with higher ambient UV, while intermittent burning exposure, a distinct exposure pattern rather than a high-dose region, increases skin injury and melanoma risk. This figure is conceptual and hypothesis-generating; it does not establish causality, an optimal UV exposure, or an individual therapeutic dose, and it requires prospective validation.

